# Twenty-Hour Hypothermic Oxygenated Perfusion Preserves Myocardial Viability and Bioenergetics with No Primary Graft Dysfunction After Porcine Heart Transplantation

**DOI:** 10.64898/2026.10.29.26366672

**Authors:** William Mitchell, Cary Karcher, Nathan Ott, Ian Mason, Diana Albino, Arianna Adamo, Fady Soliman, Friederike I. Schoettler, Vivian Zhang, Emre Bektik, Senay Ustunel, Kevin Clerkin, Christine Chan, Engel Monsanto, Miroslav Sekulic, Shaheer K. Faruqi, Craig J. Goergen, Melissa Tamimi, Morgan Moroi, Adham Elmously, Koji Takeda, Giovanni Ferrari

## Abstract

**Background:** Hypothermic oxygenated perfusion (HOPE) may extend donor heart preservation beyond conventional storage durations. We evaluated early graft function and myocardial cellular and metabolic integrity following porcine heart transplantation after 20 hours of HOPE.

**Methods:** Ten donor hearts underwent various preservation conditions: static cold storage (SCS) for 4 hours (n=2), HOPE for 4 hours (n=3), and HOPE for 20 hours (n=5), followed by heart transplantation (HT) in recipient pigs. Graft function was monitored over 3 hours by serial echocardiography, invasive hemodynamics, optical flow analysis, and vasoactive support scoring; myocardial tissue was analyzed by histopathology, flow cytometry, mitochondrial respirometry, transcriptomics, and proteomics.

**Results:** Ejection fraction, right atrial pressure and systolic blood pressure did not differ between groups, and no graft met criteria for primary graft dysfunction. LV contractility was within normal range with comparable vasoactive support among all groups. Preservation-associated weight gain was greatest after HOPE 20h, and histopathology showed comparable findings with edema present in all cohorts, without fibrosis, thrombosis, or necrosis. Cardiomyocyte viability was preserved across all groups. Maximal oxygen consumption rate was elevated in HOPE 20h versus HOPE 4h and SCS 4h, while basal and non-mitochondrial respiration were similar. Transcriptomics and proteomics revealed limited differences in gene and pathway expression among groups.

**Conclusions:** HOPE preserves donor heart viability, mitochondrial integrity, and ventricular function after 20 hours of preservation, without evidence of primary graft dysfunction, supporting extended preservation for heart transplantation. Edema, recovery of function, and metabolic responses warrant further investigation with longer-term monitoring.

Graphical Abstract

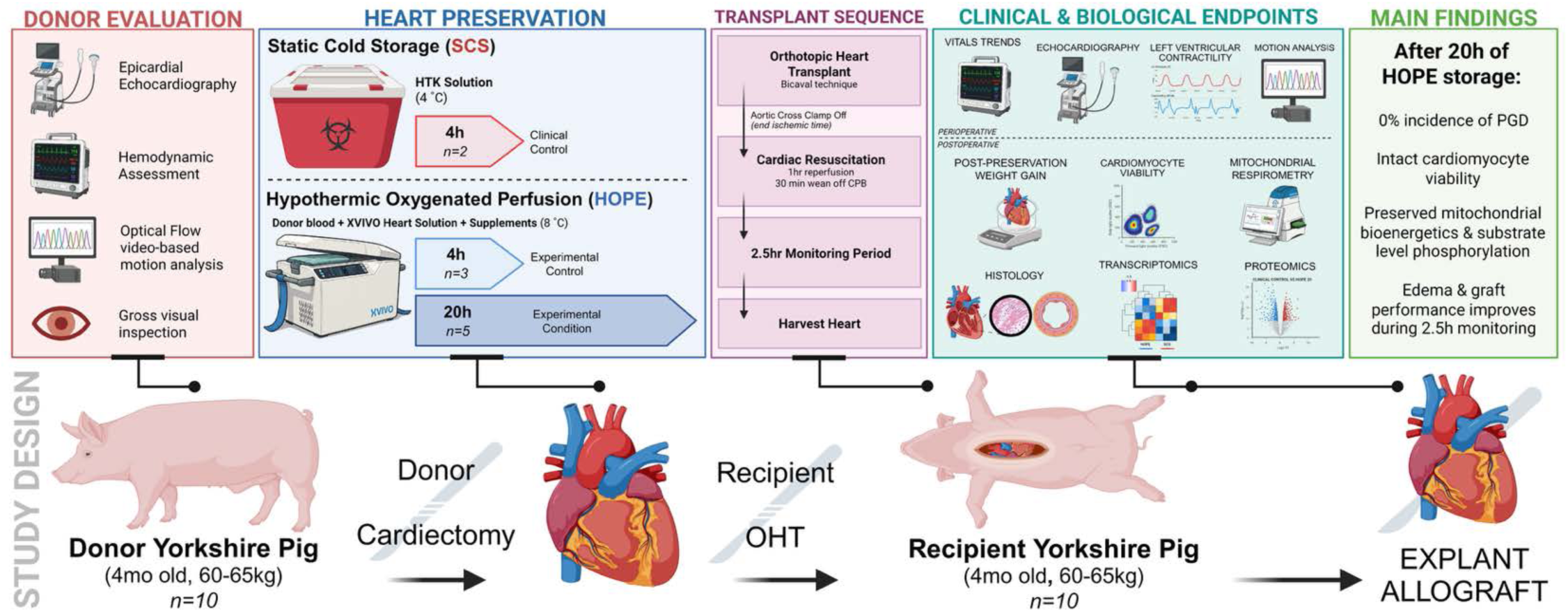

## Introduction

Heart transplantation (HT) provides the greatest clinical improvement and quality-of-life benefit for patients with end-stage heart failure beyond medical therapy and mechanical circulatory support (MCS), yet the supply of suitable donor hearts remains insufficient.^1–4^ Expanded donation criteria have increased the donor pool, but at the cost of more high-risk grafts, a risk that must be weighed against waitlist morbidity and mortality.^5–7^ Geographic reach is also limited by the maximum tolerated ischemic time of roughly 4-6 hours for static cold storage (SCS), the current clinical standard, beyond which early graft dysfunction, rejection, and poor outcomes become more likely.^8–10^ Improved preservation technology could extend this window and enable recovery of high-risk grafts.^11–13^

Hypothermic machine perfusion (HOPE) has emerged as a novel storage strategy that may safely extend the tolerated storage time.^14^ HOPE supplies continuous oxygen and nutrient delivery to the heart while maintaining a cold, arrested state using a solution composed of donor blood and cardioplegia perfused through an aortic root cannula. The ‘XVIVO Heart Assist Transport’ (XVIVO Perfusion, Gothenburg, Sweden) is among the HOPE devices currently undergoing clinical trials.^15–17^ Early trials suggest that HOPE may lower incidence of graft failure through reduced oxidative stress and more robust mitochondrial protection.^18–20^ However, the cellular and molecular changes during and after non-ischemic heart preservation remain incompletely characterized. ^12,19,20^

Our group previously demonstrated recovery of porcine hearts following up to 48 hours of HOPE preservation using an ex-vivo normothermic machine perfusion (NMP) circuit. Hearts achieved reanimation with coordinated cardiac contraction and conservation of cardiomyocyte viability.^19^ Complementary transcriptomic and metabolomic analyses supported the maintenance of molecular programs associated with myocardial viability and metabolic stability during extended preservation, including limited activation of inflammatory and injury-related pathways. We also independently identified myocardial edema as a consistent feature of HOPE preservation following both brain death (DBD) and circulatory death (DCD) procurement, with more pronounced interstitial edema following prolonged HOPE.^19^

Large animal models have demonstrated that the upper limit for HOPE may far exceed the 4-6 hour constraints of SCS while maintaining satisfactory outcomes. Thomassen et al. recently demonstrated successful porcine DBD and DCD heart transplantation following 24 hours HOPE, with preserved biventricular function, adequate cardiac output, and no primary graft dysfunction during a two-hour post-bypass observation period.^21^ Building on these findings, we provide the first functional assessment of integrated cellular, molecular, and bioenergetic characterization of heart allografts following extended HOPE, combining assessment of myocardial cell viability, transcriptomic and proteomic responses, and mitochondrial respiratory function with evaluation of early graft performance after 20 hours of preservation.

## Materials and Methods

### Animal Model and Study Design

Orthotopic heart transplantation (bicaval technique) was performed in 4-month-old female Yorkshire pigs (60-65 kg). Donor hearts were procured by direct procurement, as previously described, and preserved by static cold storage (SCS; HTK solution, 4h, n=2) or hypothermic oxygenated perfusion (HOPE; XVIVO Heart Assist Transport) for 4h (n=3) or 20h (n=5) before implantation. ^19,20^ The protocol was approved by the Columbia University Institutional Animal Care and Use Committee (AC-AACA3703). Anesthesia, procurement, cardioplegia, and implantation details are in the Supplementary Methods.

### Recipient Implant and Postoperative Monitoring

After native cardiectomy, donor hearts were implanted using bicaval technique. Animals were maintained on full CPB for 1 hour after reperfusion, then weaned over 30 minutes, with CPB discontinued at 1.5 hours. The chest was left open and animals were monitored for 3 hours after separation from CPB, with hemodynamics and vasoactive support recorded every 15 minutes, and arterial blood gases and serum chemistries hourly. Anesthetic, hemodynamic, arrhythmia, and ventilator management are described in the Supplementary Methods.

### Clinical Endpoints

The primary outcome was primary graft dysfunction (PGD), defined as: (1) failure to separate from cardiopulmonary bypass (CPB) with adequate hemodynamics after a 1.5-hour rest and weaning protocol; (2) LVEF <40% with persistently insufficient hemodynamics despite high-dose inotropes; or (3) right atrial pressure (RAP) >15 mmHg. Graft function was assessed by echocardiography (donor baseline; 0.5h, 1.5h, and 2.5h off CPB), LV contractility by solid-state pressure catheter and a composite vasoactive-inotropic drip score (1 and 2 h off CPB), and epicardial motion by optical flow analysis of video (baseline; 1.5h and 2.5h off CPB; Supplementary Methods).

### Tissue Collection

At the conclusion of the monitoring period, pigs were exsanguinated and hearts were excised warm without inducing cardiac arrest. Explanted allografts were systematically biopsied from the LV free wall and interventricular septum (IVS) for histology, flow cytometry, mitochondrial respirometry, RNA sequencing, and protein expression analysis.

### Histology, Flow Cytometry, and Mitochondrial Respirometry

LV and IVS tissue were fixed and stained with hematoxylin and eosin, Masson’s trichrome, and TUNEL for histological analysis. IVS tissue was enzymatically dissociated for flow cytometric assessment of cardiomyocyte viability (Calcein-AM^+^/DRAQ5^+^). LV free wall was sectioned into 150 µm precision-cut slices using a vibratome and processed for extracellular flux (Seahorse) mitochondrial respirometry using a modified Mito Stress Test (FCCP and rotenone/antimycin A, without oligomycin). Detailed protocols in the Supplementary Methods.

### Bulk RNA-seq Differential Expression and Gene Set Enrichment Analysis

Bulk RNA sequencing of IVS tissue was performed on HOPE 20h (n=4) and clinical-control (SCS 4h + HOPE 4h; n=4) samples. Differential expression was assessed with PyDESeq2 (Benjamini-Hochberg FDR <0.05, |log₂FC| >1), and genes ranked by Wald statistic were analyzed by GSEA against the MSigDB Hallmark, GO Biological Process, and Reactome collections (positive NES = enrichment in HOPE 20h; FDR <0.05). Full library preparation, sequencing, ortholog mapping, and analysis details are in the Supplementary Methods.

### Proteomics

Proteomic profiling of IVS tissue was performed by data-independent acquisition (diaPASEF) LC-MS/MS and DIA-NN. Differentially abundant proteins between HOPE 20h (n=4) and clinical controls (n=4) were identified with DIA-Analyst, and pre-ranked GSEA of the protein-level statistic used the same gene set collections as the RNA analysis (FDR <0.05). Results reflect relative protein abundance and pathway enrichment. Sample preparation, instrument settings, and search parameters are in the Supplementary Methods.

### Integrated Transcriptomics and Proteomics

RNA and protein results were matched by gene and pathway identifier. Directional concordance used a descriptive threshold of |log₂FC| >0.25 in both layers (Fig 8A), and the 50 genes with the lowest minimum FDR (<0.05 in at least one layer) were shown as a paired RNA–protein heatmap (Fig 8B, Supp. Table 3). Matched pathways were classified by whether FDR <0.05 was reached in the RNA data, the protein data, or both (Fig 8C). Selection and ranking details are in the Supplementary Methods.

### Statistical Analysis

Analyses were performed in GraphPad Prism. Continuous variables were compared across the three groups by one- or two-way ANOVA with Tukey’s post hoc test, or by Kruskal-Wallis test. Repeated blood gas measurements were analyzed with a mixed-effects model and Tukey’s test.

Cardiomyocyte viability used Welch’s t-tests for pairwise comparisons and Brown-Forsythe/Welch ANOVA with Dunnett’s test for the three-group comparison. Data are mean ± SEM unless stated otherwise; p<0.05 was significant. Omics statistics are described above and in the Supplementary Methods.

## Results

### Early Graft Function is Retained after 20-hour HOPE

Ten porcine donor hearts were procured and preserved by SCS for 4 h (n=2), HOPE for 4 h (n=3), or HOPE for 20 h (n=5), then transplanted into recipient pigs. Graft function was assessed during a 3-hour observation period after separation from CPB (**Supp. Video 1**). No graft met the criteria for primary graft dysfunction with elevated right atrial pressure (RAP) in the HOPE 20h group above other groups, though not statistically significant (**Fig 1A**). Systolic blood pressure, arterial lactate, pH, creatinine, and hematocrit did not differ significantly between SCS 4h, HOPE 4h, and HOPE 20h hearts during the monitoring period with vasoactive-inotropic support (combined drip score) only differing between SCS 4h and HOPE 4h groups (p<0.05) (**Fig 1B-G**). Recipient aortic cross clamp duration and CPB times did not differ between groups (**Fig. 1H-J**). Echocardiographic ejection fraction, fractional area change, and LVEDD did not differ significantly between groups at any timepoint, including compared to donor baseline function prior to heart procurement (**Fig. 2A-D** and **Supp. Video 2**). Following reperfusion, myocardial edema was observed in all groups with significant increase in interventricular wall diameter observed in the HOPE 20h group from 1 to 2 hours (**Fig. 2D-E**).

**Figure 1.**
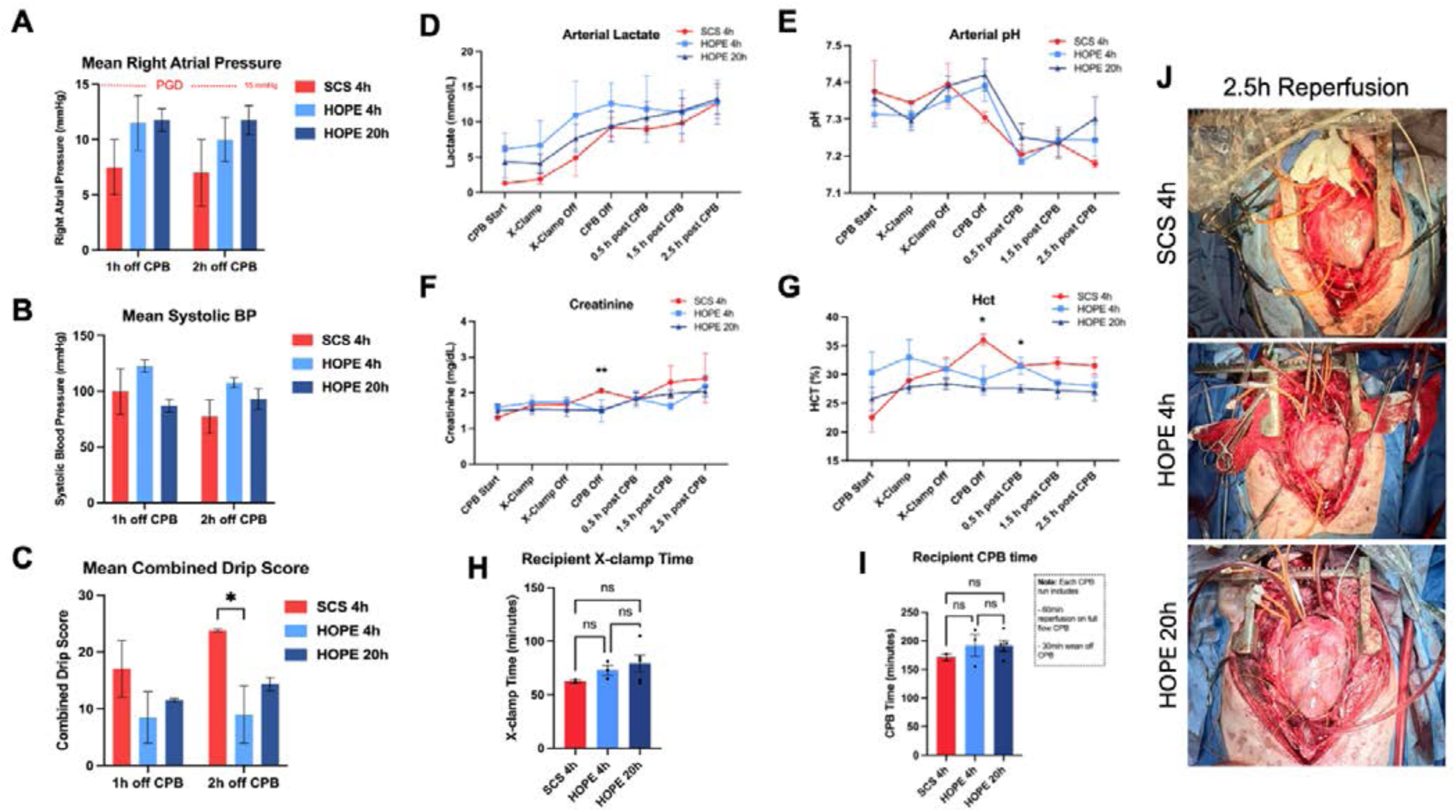
Perioperative Hemodynamics, Vasoactive Drip Score, ABG Profile, Cross-Clamp and CPB Characteristics. (A) Mean right atrial pressure (RAP) at 1 and 2 hours off cardiopulmonary bypass (CPB) across SCS 4h, HOPE 4h, and HOPE 20h groups (two-way ANOVA with Tukey’s post-hoc test). The dashed red line indicates the RAP threshold (>15 mmHg) used to indicate graft dysfunction; RAP remained below this threshold in all groups at both timepoints. (B) Mean systolic blood pressure at 1 and 2 hours off CPB. (C) Mean combined vasoactive/inotropic drip score at 1 and 2 hours off CPB (p<0.05). (D) Arterial lactate, (E) arterial pH, (F) creatinine, and (G) hematocrit measured serially from CPB initiation through 2.5 hours post-CPB. Creatinine was significantly higher in 4h SCS compared to both HOPE groups at CPB off (p<0.01). Hematocrit was significantly higher in 4h SCS compared to both HOPE groups at CPB off and at 0.5 hours post-CPB (p<0.05 at each timepoint). (H) Recipient cross clamp duration measured as time elapsed from application of ascending aortic occlusion clamp to start of reperfusion. (I) Total recipient cardiopulmonary bypass times. (J) Representative intraoperative photographs of the transplanted graft at 2.5 hours of reperfusion in SCS 4h, HOPE 4h, and HOPE 20h groups. Data in A–I are presented as mean ± SEM. SCS 4h, n=2; HOPE 4h, n=3; HOPE 20h, n=5.

**Figure 2.**
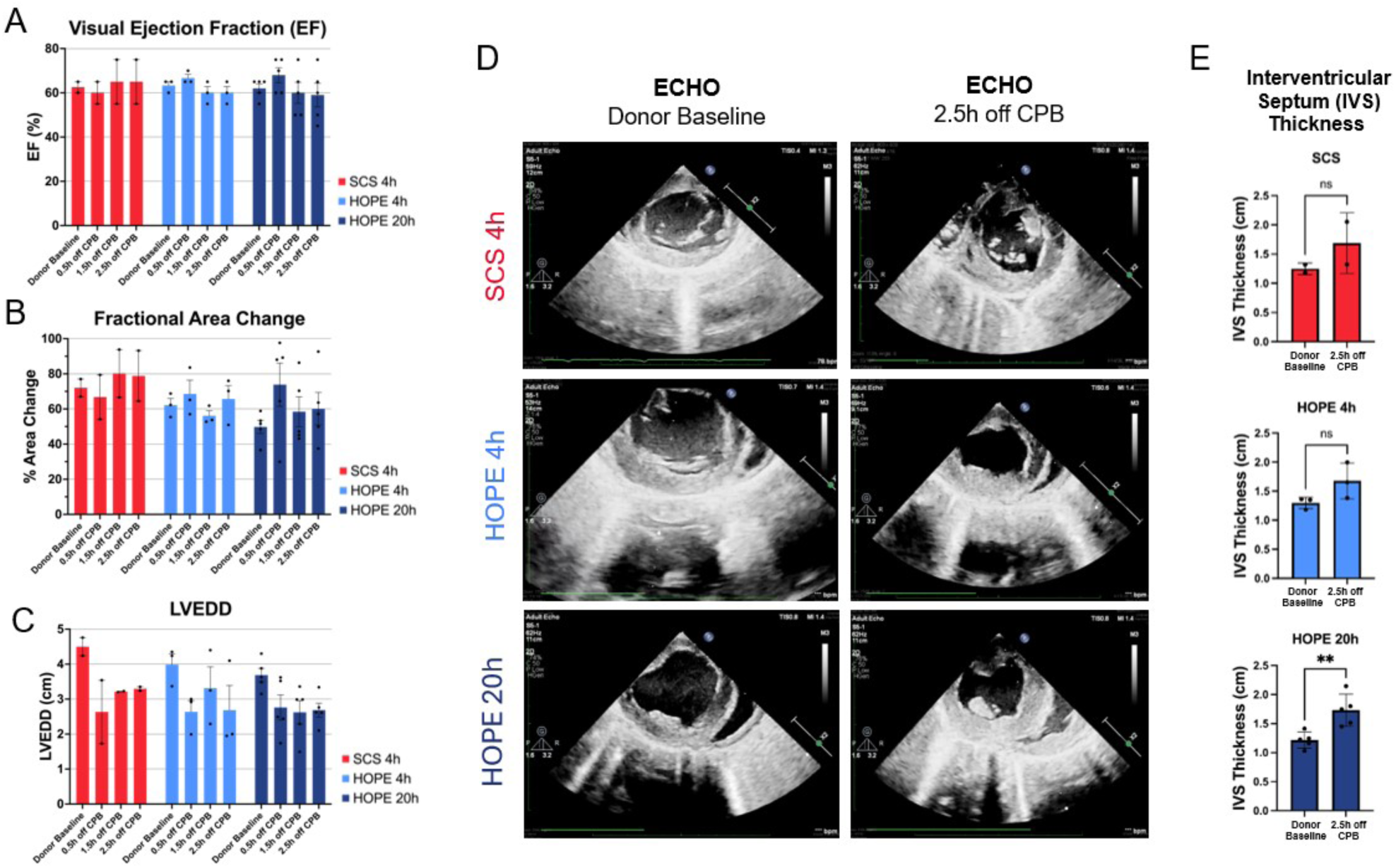
Echocardiographic Assessment of Graft Function and Ventricle Morphology. (A) Visual ejection fraction (EF), (B) fractional area change, and (C) left ventricular end-diastolic diameter (LVEDD), assessed by echocardiography at donor baseline and at 0.5, 1.5, and 2.5 hours off cardiopulmonary bypass (CPB) in SCS 4h, HOPE 4h, and HOPE 20h groups (two-way ANOVA with Tukey’s post-hoc test). Individual data points are overlaid on each bar. (D) Representative echocardiographic images at donor baseline and 2.5 hours off CPB for each preservation group. (E) Interventricular septum diameter (cm) in SCS 4h, HOPE 4h, and HOPE 20h groups compared at baseline and at 2.5h off CPB.

### Left Ventricular Contractility and Epicardial Motion Remain Within Normal Range After 20-hour HOPE

Contractile function was assessed by LV pressure-derived contractility at 1 and 2 hours off CPB, and by optical flow analysis of epicardial motion at baseline and at 1.5 and 2.5 hours off CPB (**Fig 3** and **Supp. Video 3**). LV contractility did not differ between groups at 1 hour off CPB. At 2 hours, contractility was significantly lower in HOPE 20h than in HOPE 4h (p<0.05), with no significant difference between SCS 4h and either HOPE group. All groups showed an increase in contractility from 1 to 2 hours off CPB, and all values remained within the normal range for juvenile Yorkshire pig (**Fig. 3A-B**).^22^ Peak epicardial contraction velocity did not differ significantly between groups or timepoints, including baseline (48.2 ± 17.0 mm/s), with all groups remaining comparable to baseline through 2.5 hours off CPB (**Fig. 3C-D**). Similarly, there was no significant difference in peak contraction acceleration between groups, with baseline acceleration 396.8 ± 153.7 mm/s^2^. All groups exhibited a mild, but not statistically significant, increase in peak contraction acceleration from 1.5h to 2.5h, with the HOPE 20h group showing the least increase (1.5h, 505.7 ± 281 mm/s^2^; 2.5h, 522.5 ± 239.2 mm/s^2^) (**Fig. 3E**).

**Figure 3.**
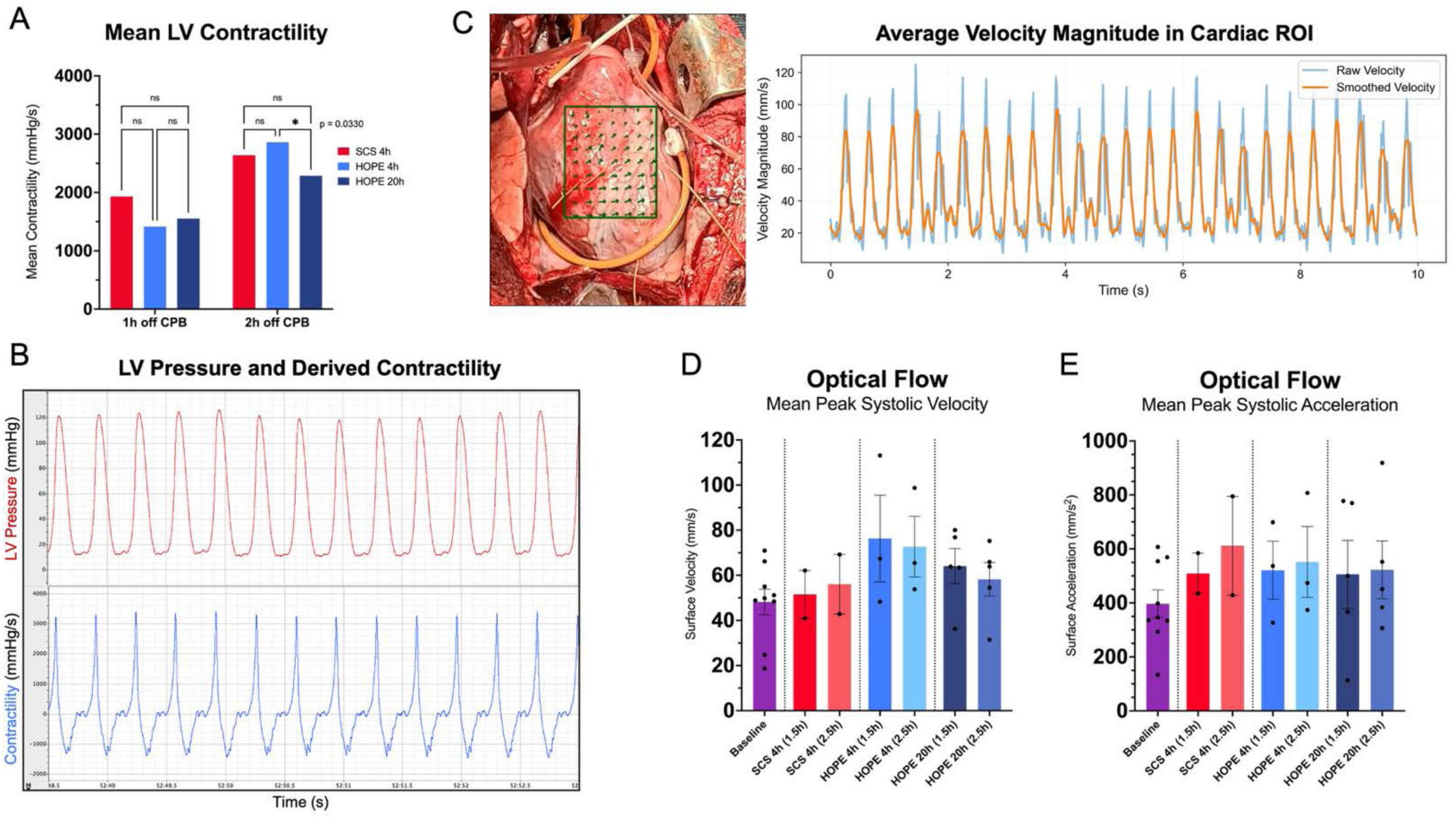
LV Pressure Derived Contractility and Epicardial Wall Motion Assessment. (A) Mean left ventricular contractility, measured via solid-state LV pressure catheter, at 1 and 2 hours off CPB across all three groups. Contractility did not differ significantly between groups at 1 hour off CPB; at 2 hours off CPB, contractility was significantly lower in HOPE 20h compared to HOPE 4h (*p<0.05), with no significant difference between SCS 4h and either HOPE group. (B) Example of LV Pressure waveform and contractility obtained from solid-state catheter. (C) Representative speckle tracking vectors assigned to anterior interventricular sulcus for video-based motion quantification (left). Average ventricular wall velocity over time with both raw and smooth velocity calculation (right). (D) Mean peak epicardial contraction velocity and (E) mean peak epicardial contraction acceleration, quantified by optical flow analysis, at donor baseline and at 1.5 and 2.5 hours off CPB. Contraction velocity and acceleration did not differ significantly between groups or timepoints. Data are presented as mean ± SEM.

### Extended HOPE is Accompanied by Greater Weight Gain and Edema

To characterize edema and structural injury after extended HOPE, we assessed gross tissue morphology, histopathology, pathways related to vascular permeability, and heart weight gain (**Fig 4**). All hearts had preserved gross morphology without visible necrosis (**Fig 4A**). GSEA of vascular permeability and edema pathways showed significant enrichment of gap junction trafficking and endothelial cell migration at the RNA level and of cell-cell junction organization at the protein level, whereas endothelial barrier, aquaporin-mediated transport, and VEGFR2-mediated permeability pathways were not significant (**Fig 4B**). Blinded histopathology showed interstitial edema in all samples, most pronounced in HOPE 20h. Coagulative myocyte necrosis was focal; acute inflammation and hemorrhage were confined to one HOPE 20h animal, and no fibrosis or thrombosis was observed. TUNEL staining showed rare apoptotic nuclei without differences between groups (**Fig 4C**, **Supplemental Fig 1**). HOPE coronary flow (mL/min) tracing over time during individual preservations with mean and +/- 1 SD demonstrates steady flow decline across all HOPE groups (**Fig 4D**). Percent heart weight gain following preservation, measured before implantation, was minimal in SCS 4h, intermediate in HOPE 4h, and most pronounced in HOPE 20h, which differed significantly from SCS 4h (p<0.05) but not from HOPE 4h (**Fig 4E**). Because coronary flow declined progressively during HOPE preservation, we examined the left anterior descending (LAD) coronary artery for vessel wall layer swelling that may contribute to coronary resistance. Alcian blue-stained sections showed a non-significant trend toward greater tunica intima thickness relative to tunica media after extended HOPE (**Fig 4F-G**).

**Figure 4.**
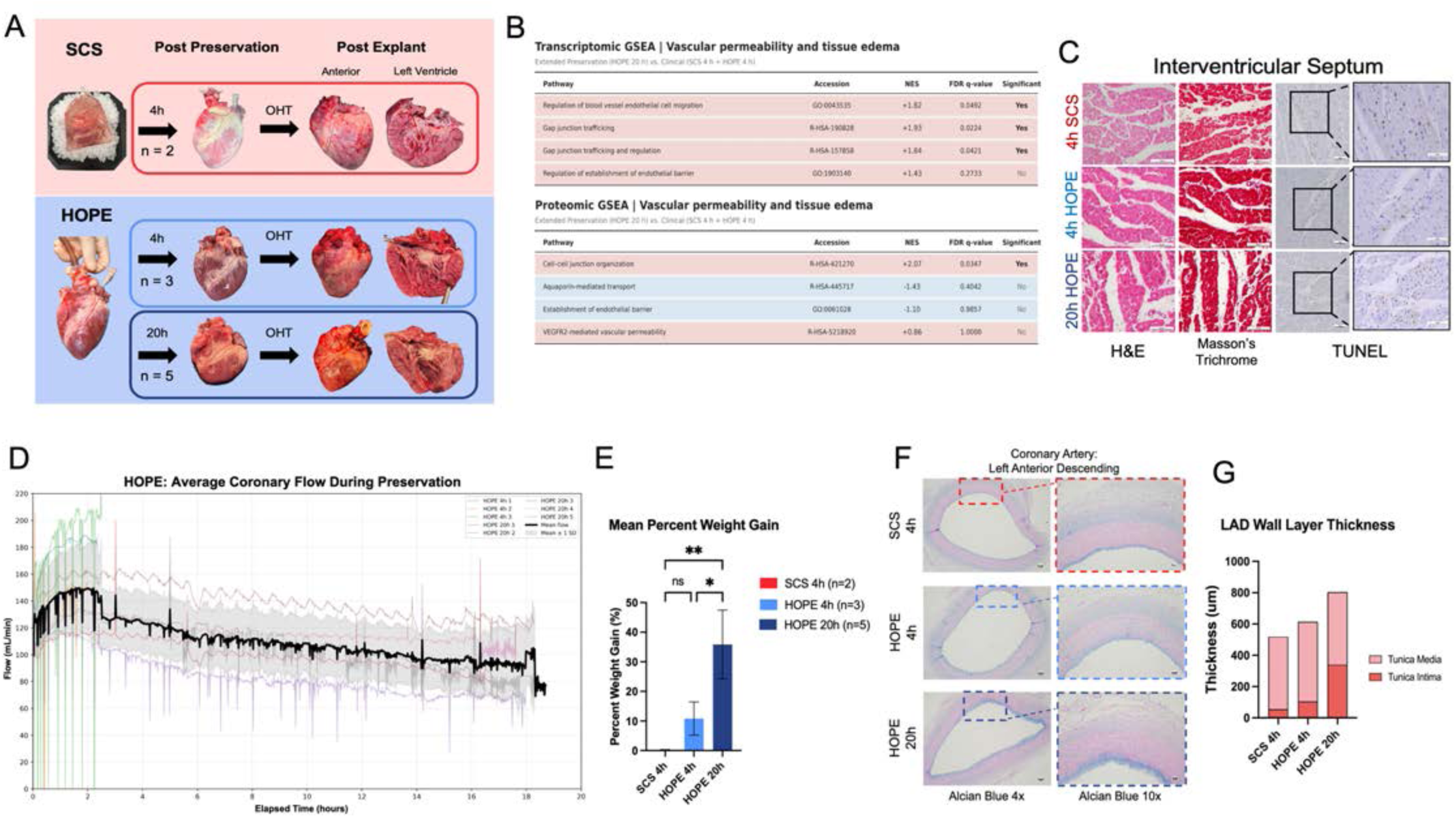
Explant Gross Anatomy, Histopathology, and Weight-Based Assessment. (A) Representative gross images of hearts from the SCS (n=2) and HOPE (4h n=3 and 20h n=5) groups at donor procurement, following preservation, and following explant HT, shown in anterior and left ventricular cross-sectional views. (B) Transcriptomic GSEA: Significant positive enrichment was identified for regulation of blood vessel endothelial cell migration (NES +1.82), gap junction trafficking (NES +1.93), and gap junction trafficking and regulation (NES +1.84). Regulation of establishment of endothelial barrier demonstrated positive enrichment (NES +1.43) but did not meet the FDR <0.05 significance threshold. Proteomic GSEA: Significant positive enrichment was identified for cell–cell junction organization (NES +2.07). Aquaporin-mediated transport (NES −1.43), establishment of endothelial barrier (NES −1.10), and VEGFR2-mediated vascular permeability (NES +0.86) did not meet the FDR <0.05 significance threshold. (C) Representative hematoxylin and eosin (H&E), Masson’s trichrome, and TUNEL-stained sections of the interventricular septum from SCS 4h, HOPE 4h, and HOPE 20h hearts taken at 40x magnification. (D) Average coronary flow during HOPE preservation from both 4h and 20h groups (black line) with individual flow curves superimposed in the background (E) Percent weight gain following preservation in SCS 4h (n=2), HOPE 4h (n=3), and HOPE 20h (n=4) hearts. (F) Representative alcian blue (2.5 pH) stained sections of left anterior descending (LAD) branch of coronary artery from SCS 4h, HOPE 4h, and HOPE 20h hearts taken at 4x and 10x magnification. (G) LAD wall layer thickness in micrometers of tunica intima and tunica media across all groups SCS 4h (n=2), HOPE 4h (n=3), and HOPE 20h (n=4). Data are presented as mean values, no comparison reached significance.

### Myocardial Cell Viability Is Retained After Extended HOPE

To assess whether extended HOPE preserved cellular viability, interventricular septal biopsies collected at the end of the post-implantation observation period were dissociated and analyzed by flow cytometry for viable (Calcein-AM⁺/DRAQ5⁺) cells (**Fig 5A**). Viable cells were recovered from all groups, with mean viability of 86% in HOPE 20h, 81% in HOPE 4h, and 77% in SCS 4h, and no statistically significant differences between groups (p>0.05) (**Fig 5B-E**). Thus, neither 4-hour nor 20-hour HOPE reduced cardiomyocyte viability relative to the clinical-standard SCS 4h group, despite the longer preservation time and different preservation conditions.

**Figure 5:**
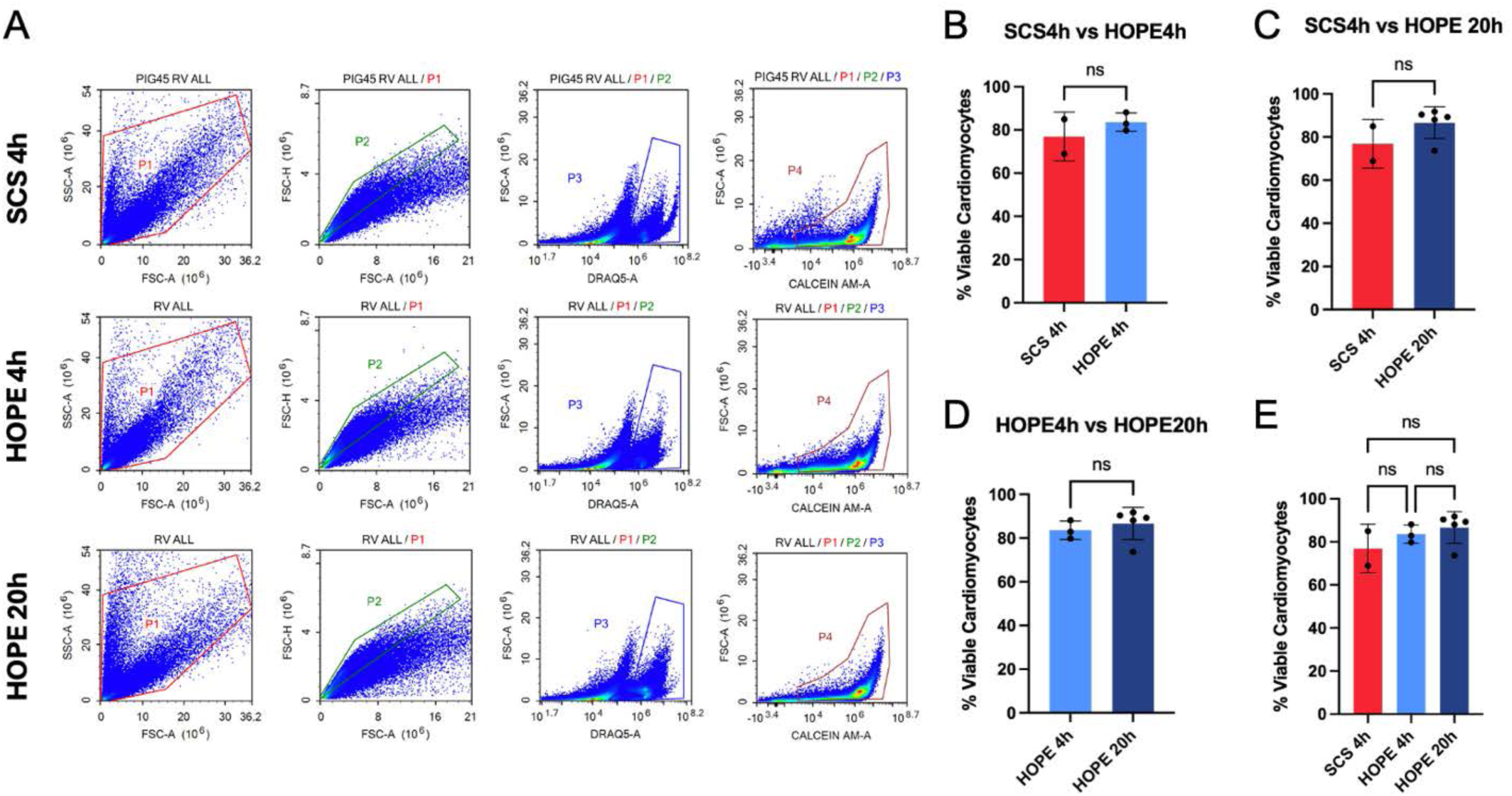
Isolation and Flow Cytometry of Viable Cardiomyocytes. (A) Representative flow cytometry plots. Events were first gated to exclude debris and doublets (P1, P2), then gated to isolate DRAQ5-positive nucleated events (P3), and finally Calcein-AM-positive metabolically active cells (P4). (B–E) Quantification of viable cardiomyocytes (% viable, Calcein-AM⁺/DRAQ5⁺) comparing (B) SCS 4h vs. HOPE 4h, (C) SCS 4h vs. HOPE 20h, (D) HOPE 4h vs. HOPE 20h, and (E) all three groups together. Data are presented as mean ± SD for SCS 4h (n=2), HOPE 4h (n=3), and HOPE 20h (n=5). Statistical analysis was performed using Unpaired t test with Welch’s correction for pairwise comparisons (B-D), and Brown–Forsythe and Welch’s one-way ANOVA followed by Dunnett’s multiple-comparisons test for the three-group comparison (E). No statistically significant differences were observed between groups (p > 0.05).

### Extended HOPE is Associated with Preserved Bioenergetics with Elevated Maximal Oxygen Consumption Rate, Spare Aerobic Respiratory Capacity and Substrate Level Phosphorylation

LV free wall tissue sections underwent extracellular flux Mito Stress Test to quantify OCR and ECAR (**Fig 6A**, **6E**). Because of the small group sizes, SCS 4h and HOPE 4h hearts were pooled as a clinical-standard comparator (n=4) and compared with HOPE 20h (n=4) (**Fig 6B**, **6F**). Basal oxygen consumption rate (OCR) did not differ between the pooled clinical group and HOPE 20h (**Fig. 6C**). Maximal OCR was significantly higher in HOPE 20h (p<0.01), while non-mitochondrial OCR remained low and did not differ significantly between groups. Spare respiratory capacity was also significantly higher in HOPE 20h (p<0.01) (**Fig. 6D**). Extracellular acidification rate (ECAR) provides a measurement to track substrate-level phosphorylation indirectly by measuring proton pumping. ECAR tracing followed a similar pattern: basal ECAR did not differ between groups, compensatory ECAR was significantly higher in HOPE 20h than the pooled clinical group (p<0.05), and non-mitochondrial ECAR did not differ significantly between groups (**Fig. 6G**).

**Figure 6.**
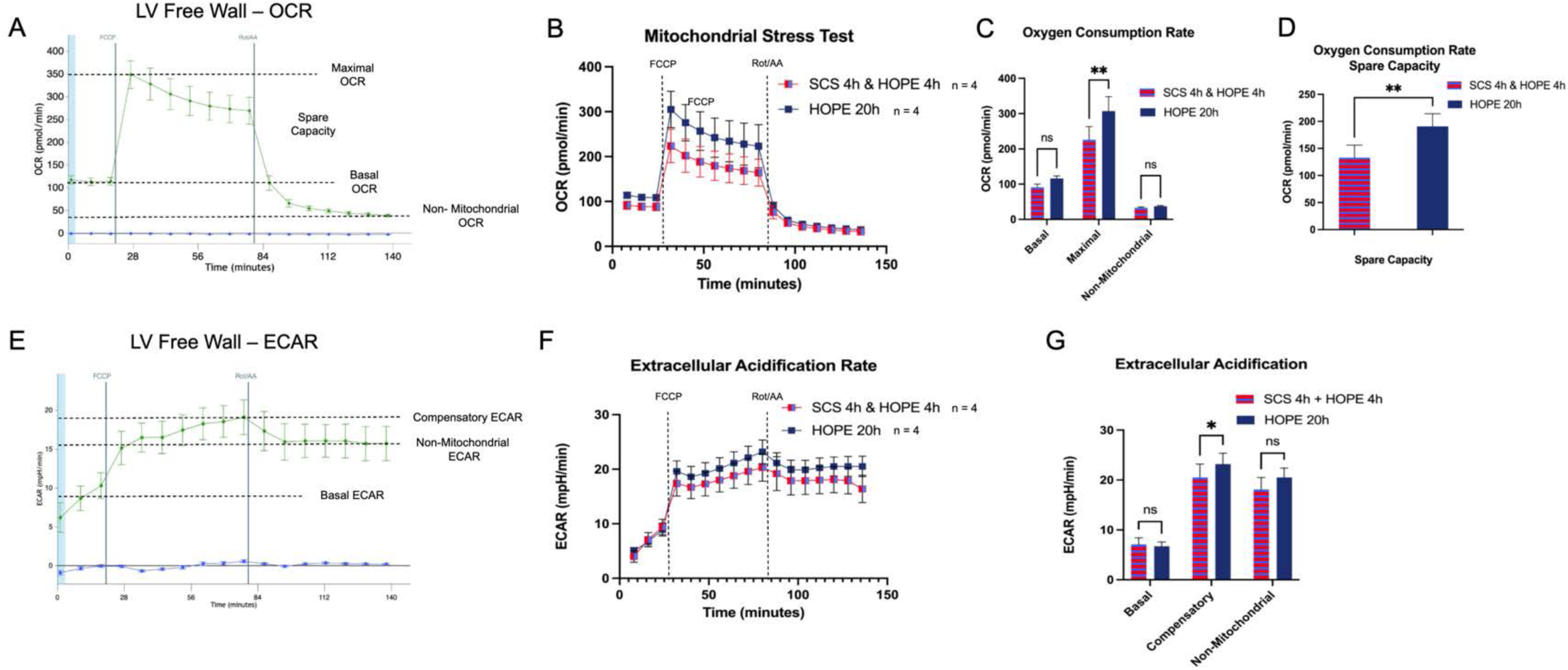
Bioenergetic tracing of mitochondrial oxidative and substrate-level phosphorylation in clinical versus extended HOPE preservation. (A) Representative oxygen consumption rate (OCR) trace of LV free wall tissue during a modified Seahorse Mito Stress Test, illustrating calculation of basal OCR, maximal OCR, spare respiratory capacity, and non-mitochondrial OCR following sequential injection of FCCP and rotenone/antimycin A (Rot/AA). (B) OCR traces comparing pooled clinical-standard preservation (SCS 4h + HOPE 4h, n=4) versus extended HOPE 20h preservation (n=4). (C) Quantification of basal, maximal, and non-mitochondrial OCR. Maximal OCR was significantly higher in HOPE 20h compared to the pooled clinical group (**p<0.01); basal and non-mitochondrial OCR did not differ significantly. (D) Spare respiratory capacity (maximal OCR − basal OCR) was significantly greater in HOPE 20h compared to the pooled clinical group (**p<0.01). (E) Representative extracellular acidification rate (ECAR) trace illustrating calculation of basal ECAR, FCCP-induced compensatory ECAR, and non-mitochondrial ECAR. (F) ECAR traces comparing pooled clinical-standard preservation versus HOPE 20h. (G) Quantification of basal, compensatory, and non-mitochondrial ECAR. Compensatory ECAR was significantly higher in HOPE 20h compared to the pooled clinical group (*p<0.05); basal and non-mitochondrial ECAR did not differ significantly. Data are presented as mean ± SEM. SCS 4h + HOPE 4h, n=4; HOPE 20h, n=4.

### Transcriptomic and Proteomic Profiling Identifies Distinct Molecular Responses to Extended Hypothermic Oxygenated Heart Preservation

To characterize molecular responses to extended preservation, bulk RNA sequencing and proteomics were performed on interventricular septal biopsies collected at the end of the observation period, comparing HOPE 20h with pooled clinical controls (SCS 4h and HOPE 4h). RNA sequencing identified 68 differentially expressed genes (DEGs; FDR <0.05, |log₂FC| >1; **Supplemental Table 1**) reflecting inflammation, cellular stress response, and metabolism (**Fig 7A-B**). Gene set enrichment analysis (GSEA) showed positive enrichment of cellular stress response, RNA processing, and extracellular matrix remodeling and negative enrichment of fatty acid metabolism (**Fig 7C; Supp. Figs 3-6**). In order to understand whether specific genes drove GSEA results, leading edge analysis was performed and when controlling for pathway redundancy, leading edge genes were consistently present in functionally distinct pathway clusters; informing potential central drivers of cellular response to extended preservation, these genes may highlight future directions (**Supp. Fig 7**). Proteomics identified 39 differentially abundant proteins (29 higher and 10 lower in HOPE 20h; **Fig 7D-E**, **Supp. Table 2**), with less concordance to the transcriptomic findings, possibly reflecting a lag between transcript and protein changes within the short observation period. Proteomic GSEA showed positive enrichment of neutrophil migration and mitochondrial translation and negative enrichment of coagulation, complement cascade, and platelet degranulation (**Fig 7F; Supp. Fig 8**).

**Figure 7.**
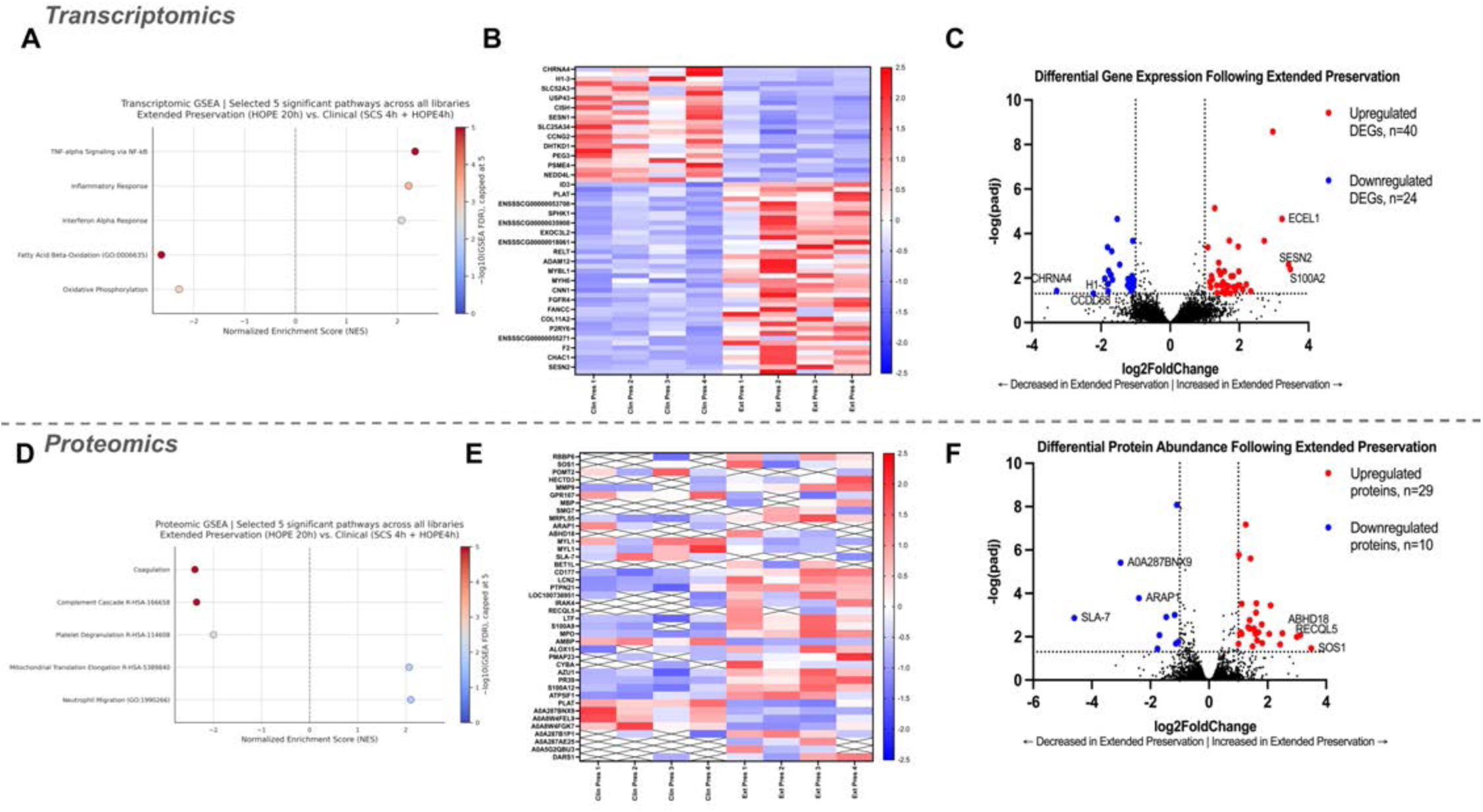
Differential Transcriptomic and Proteomic Profiles Following Extended HOPE. **(A)** Gene set enrichment analysis (GSEA) of transcriptomic data illustrating five selected significantly enriched pathways across Gene Ontology (GO), Hallmark, and Reactome gene set libraries. **(B)** Heatmap displaying standardized transcriptomic expression profiles across individual preservation samples (clinical controls, n=4; HOPE20h, n=4). 30 representative differentially expressed genes visualized. **(C)** Volcano plot illustrating differential gene expression between HOPE20h and Clinical Controls. Red indicates significantly upregulated genes (n=40), blue indicates significantly downregulated genes (n=24), and black indicates genes not meeting the displayed significance and fold-change thresholds. Vertical dotted lines indicate absolute log₂FC thresholds of 1, and the horizontal dotted line indicates -log(0.05). Top 3 most differentially upregulated and downregulated by log₂FC are annotated. **(D)** GSEA of proteomic data illustrating five selected significantly enriched pathways across GO, Hallmark, and Reactome libraries. **(E)** Heatmap displaying standardized protein abundance across individual preservation samples (clinical controls, n=4; HOPE20h, n=4). **(F)** Volcano plot illustrating differential protein abundance between HOPE20h and clinical controls. Top 3 most differentially upregulated and downregulated by log₂FC are annotated.

### Integrated Multi-Omics Analysis Describes Heterogeneous RNA–Protein Concordance and Predominant Transcriptomic Pathway Enrichment

To relate transcriptional changes to protein abundance, we integrated the bulk RNA-seq and DIA-MS datasets by matching genes and pathways. Of 656 matched RNA–protein pairs exceeding a descriptive threshold of |log₂FC| >0.25 in both layers, 419 (64%) were directionally concordant and 237 (36%) discordant; 4,966 pairs did not meet the threshold (**Fig 8A**). At the gene level, only one concordant pair, LIPE decreased in HOPE 20h, reached FDR <0.05 in both layers (**Fig 8B, Supp. Table 3**), indicating limited RNA–protein coordination. At the pathway level (**Fig 8C**), the pathways meeting FDR <0.05 in at least one layer were those identified by transcriptomics, with strong positive enrichment of RNA processing and negative enrichment of fatty acid β-oxidation. These pathways did not reach FDR <0.05 in the proteomic data, but most showed the same direction of change. We also scored curated cardiac cell-type marker panels in the bulk RNA-seq data (**Supp. Fig 9A**). SCS 4h and HOPE 4h samples did not separate by cell-type signature (**Supp. Fig 9B**). Relative to pooled controls, HOPE 20h showed higher signature scores for B cells, T cells, macrophages, fibroblasts, and vascular smooth muscle cells and lower scores for endothelial cells and pericytes (**Supp. Fig 9C**). These shifts are directionally consistent with the inflammatory and vascular-permeability signals described above (**Fig 4B**).

**Figure 8.**
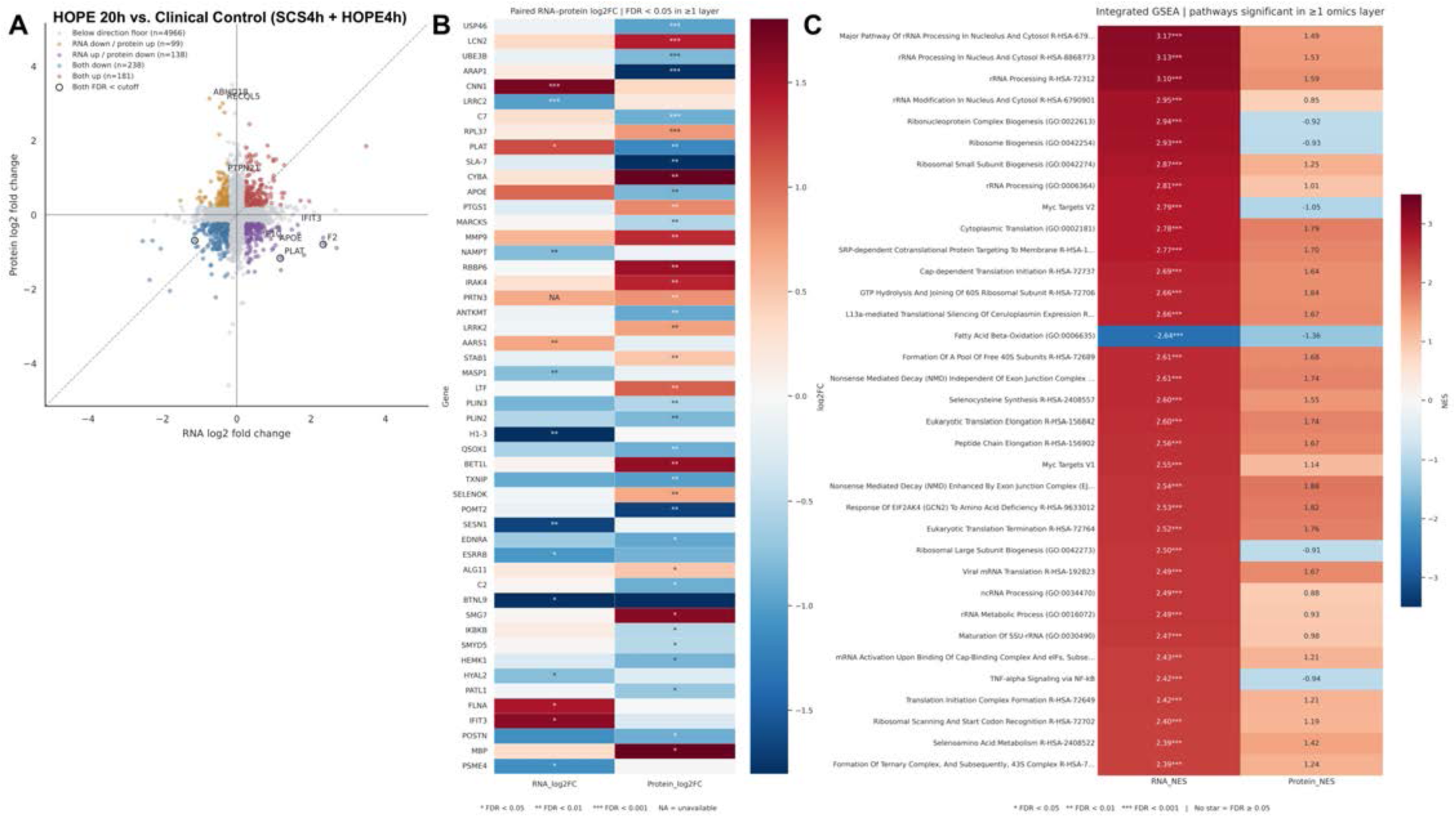
Integrated Transcriptomic and Proteomic Analysis Following Extended HOPE. (A) Scatterplot illustrating paired RNA and protein log₂ fold changes (log₂FC) for genes mapped across both omics datasets. Each point represents a matched gene–protein pair, with RNA log₂FC on the horizontal axis and protein log₂FC on the vertical axis. The diagonal reference line represents equivalent RNA and protein log₂FC values. Outlined points identify pairs meeting the false discovery rate (FDR) <0.05 threshold in both omics layers. (B) Concordance heatmap of the 50 highest-ranked RNA-Protein pairs by minimum FDR where at least 1-member of the pair is FDR <0.05. Asterisks indicate statistical significance within each omics layer (*FDR <0.05; **FDR <0.01; ***FDR <0.001). (C) Integrated gene set enrichment analysis (GSEA) comparing normalized enrichment scores (NES) between transcriptomic and proteomic datasets for pathways meeting FDR <0.05 in at least one omics layer, asterisks denoting statistical significance within the corresponding omics layer (*FDR <0.05; **FDR <0.01; ***FDR <0.001).

## Discussion

In this study, 20-hour hypothermic oxygenated perfusion supported successful porcine heart transplantation: all hearts were weaned from cardiopulmonary bypass, and none met study-defined criteria for primary graft dysfunction during the three-hour observation period. Ejection fraction and systemic hemodynamics did not differ between groups, and LV contractility, although lower after HOPE 20h than HOPE 4h at two hours, remained within the normal range. Extended HOPE showed cell viability comparable to clinical controls and preserved mitochondrial respiratory capacity, with distinct transcriptomic and proteomic changes in inflammatory signaling, RNA processing, and energy metabolism and limited RNA-protein concordance. It was also accompanied by heart weight gain and interstitial edema. Edema remains one of the main barriers to clinical translation and a priority for mitigation and longer postoperative monitoring studies.

Retained myocardial cell viability after 20h HOPE complements the observed early graft recovery and extends our previous benchtop findings into *in vivo* heart transplant model. The elevated maximal oxygen consumption rate (OCR) observed in the HOPE 20h group likely reflects a compensatory mitochondrial response to the cumulative stress of prolonged ischemia and reperfusion, rather than a marker of more robust tissue health.^23,24^ In separate work applying this same Seahorse respirometry assay to grossly necrotic, non-viable myocardium, we did not observe this compensatory elevation in OCR or ECAR; rather, energy production in severely injured tissue was uniformly and substantially depressed, with no comparable rise in any parameter (**Supplemental Fig. 2**). This suggests that the elevated maximal OCR seen in HOPE 20h hearts represent still-viable myocardium actively performing oxidative metabolism to compensate for and repair ischemic-reperfusion injury, rather than a failing or irreversibly injured tissue bed.

Our data, and others, suggest that hearts preserved with HOPE for up to 20 hours retain sufficient function to undergo transplantation.^20^ However, the intermediate and long-term consequences of the observed diastolic dysfunction and impaired lusitropy in the setting of pronounced myocardial edema are unknown. Importantly, edema and diastolic dysfunction were also observed in our SCS cohort and are frequently observed in current practice. Though some amount of edema and diastolic dysfunction are inherent to both heart transplantation, and native to a porcine model, it is highly likely HOPE is a separate contributing factor to interstitial edema.^24,25^ The continuous perfusion of the graft with preservation solution may contribute to the edema and weight gain observed after HOPE preservations. Additionally, we observed a reduction in perfusion rate as preservation time elapsed. Therefore, we hypothesize there is a component of coronary vasoconstriction and/or myocardial stiffness that contributes to increased vascular resistance, resulting in reduced flow. Mitigating this edema-associated diastolic impairment by targeting the underlying mechanisms represents an important and clinically actionable next step for future studies aimed at optimizing and extending the HOPE preservation platform.

Several limitations of this study warrant discussion. The small, unequal cohorts, particularly the SCS 4h group (n=2), compared with HOPE 4h (n=3) and HOPE 20h (n=5), limit statistical power, and nonsignificant differences should not be interpreted as equivalence. Larger experimental cohorts are needed to strengthen these comparisons. Additionally, this study did not include long-term survival data; animals were not recovered to an intensive care setting, and the longer-term functional trajectory and recovery potential of hearts preserved for 20 hours with HOPE remains presently unknown.

## Acknowledgments

Histology Core: Histological specimen preparation and staining for this work were performed in the Molecular Pathology Shared Resource of the Herbert Irving Comprehensive Cancer Center at Columbia University. Seahorse XFe96 Analyzer: Extracellular flux mitochondrial stress test was performed in the Columbia Medicine Microscopy Core (MMC). This work was supported by the National Institutes of Health grant #P30 CA013696 (National Cancer Institute). JP Sulzberger Columbia Genome Center: “This research was funded in part through the National Institutes of Health/National Cancer Institute Cancer Center Support Grant P30CA013696 and used the Genomics and High Throughput Screening Shared Resource.” Columbia Stem Cell Initiative Flow Cytometry core. Columbia Biomarkers Core Laboratory.

## Financial Disclosure Statement

National Heart, Lung and Blood Institute of the National Institutes of Health (R01 HL 170573 and R01 HL 163085) (GF), Research Agreement with XVIVO Perfusion Inc. (GF) Andrew Sabin Family Foundation Cardiovascular Research Laboratory (GF).

## Conflict of Interest Statement

This work was supported in part by a research agreement between Columbia University and XVIVO Perfusion Inc. (Gothenburg, Sweden) with Dr. Giovanni Ferrari as principal investigator. Columbia University received an industry-sponsored grant from XVIVO Perfusion Inc. XVIVO also provided the Heart Assist Transport device and the perfusion consumables. XVIVO Perfusion Inc. had no role in the study design, data collection, analysis, interpretation, writing of the manuscript, or the decision to submit it for publication. The authors have no financial conflict of interest or relationships relevant to the contents of this article to disclose.

## Data Availability Statement

The RNA sequencing data discussed in this publication have been deposited in NCBI’s Gene Expression Omnibus and are accessible through GEO Series accession number GSE351179 (https://www.ncbi.nlm.nih.gov/geo/query/acc.cgi?acc=GSE351179).

## Declaration of generative AI and AI-assisted technologies

During the preparation of this work, the authors used ChatGPT, Claude.ai and Grammarly for grammatical editing and proofreading. The authors reviewed and edited the output as needed and took full responsibility for the content of the published article.

## Supplemental Videos

**Supplemental Video 1 – Transplanted Hearts, Visual Comparison (2.5h off CPB)**

https://doi.org/10.6084/m9.figshare.34614816

**Supplemental Video 2 - ECHO Comparison (2.5h off CPB)**

https://doi.org/10.6084/m9.figshare.34590096

**Supplemental Video 3 - Optical Flow Technique**

https://doi.org/10.6084/m9.figshare.34590525

---

**Supplemental Figure 1:**
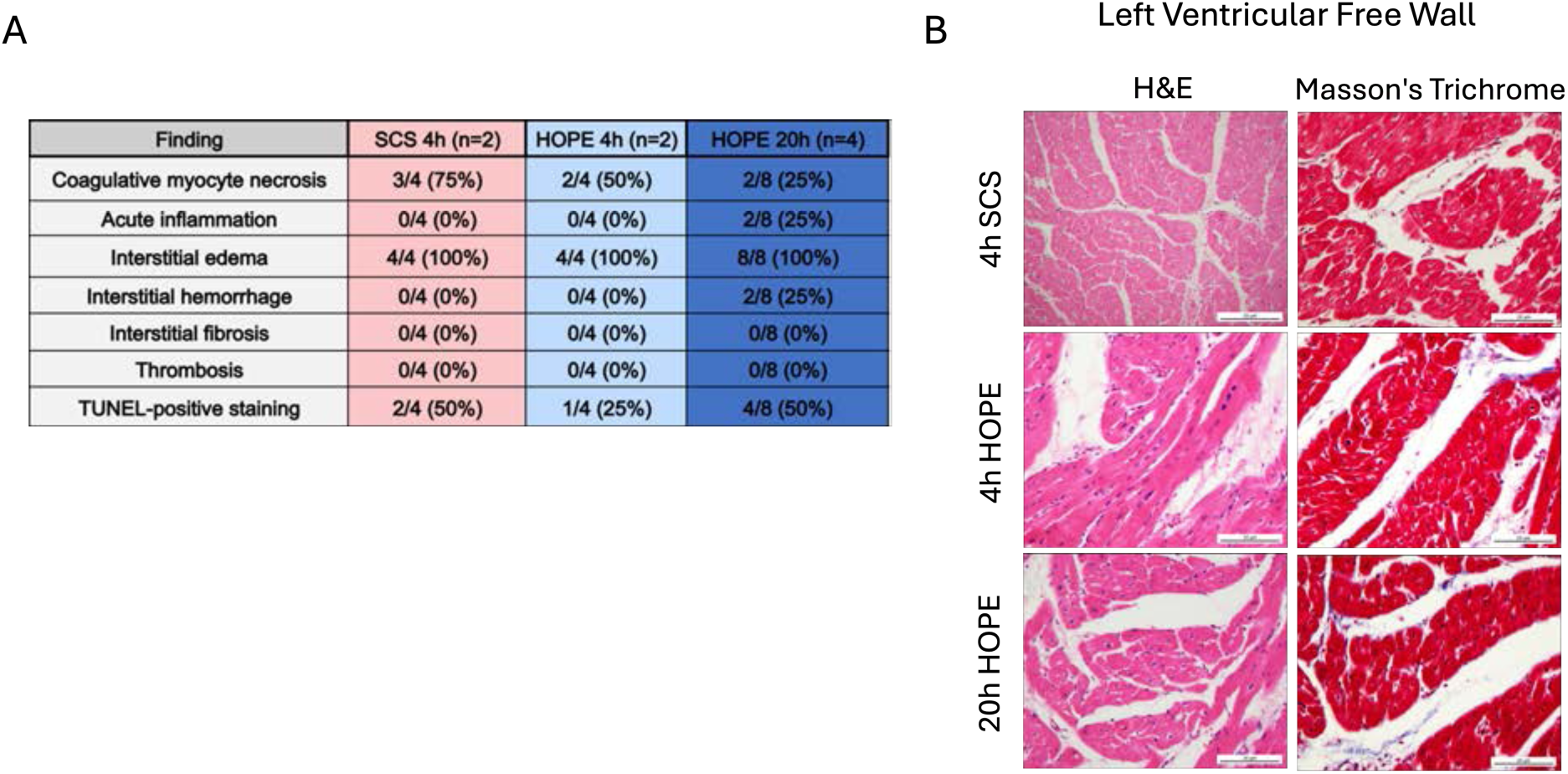
Quantitative histopathologic findings and representative left ventricular histology. (A) Summary of histopathologic findings across preservation groups, expressed as the proportion of samples positive for each finding. Coagulative myocyte necrosis decreased with increasing preservation duration (SCS 4h, 75%; HOPE 4h, 50%; HOPE 20h, 25%). Interstitial edema was present in all samples across all groups. Acute inflammation and interstitial hemorrhage were each observed in 2 of 8 HOPE 20h samples. No sample in any group demonstrated interstitial fibrosis or thrombosis. TUNEL-positive staining was identified in 50% of SCS 4h, 25% of HOPE 4h, and 50% of HOPE 20h samples. (B) Representative hematoxylin and eosin (H&E) and Masson’s trichrome-stained sections, imaged at 40x, of the left ventricular free wall from SCS 4h, HOPE 4h, and HOPE 20h hearts, demonstrating preserved myocardial architecture without significant fibrosis across all preservation groups. Scale bars, 20 µm.

**Supplemental Figure 2.**
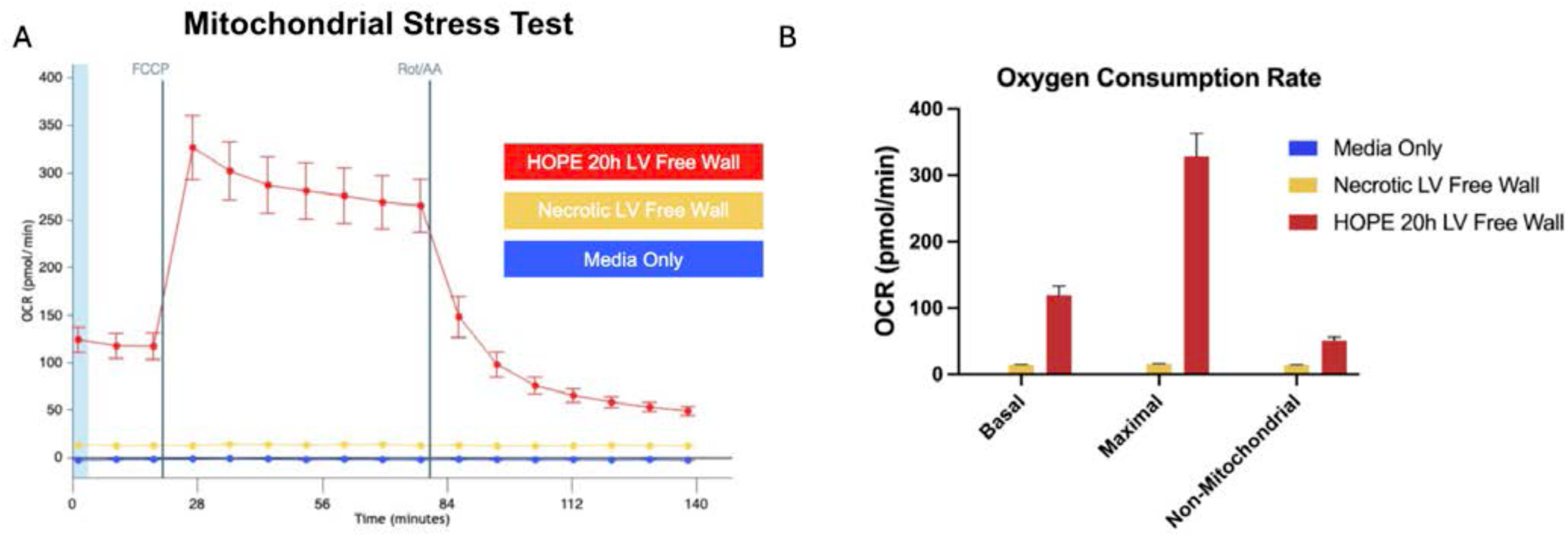
Mitochondrial Respirometry Distinguishes Viable HOPE 20h Myocardium from Necrotic Tissue. (A) Oxygen consumption rate (OCR) traces from a Seahorse Mito Stress Test comparing media-only wells (blue), necrotic left ventricular (LV) free wall tissue (yellow; explanted tissue held at room temperature for 24 hours to induce cell death), and HOPE 20h LV free wall tissue (red), with sequential injection of FCCP and rotenone/antimycin A (Rot/AA). (B) Quantification of basal, maximal, and non-mitochondrial OCR across the three conditions. Necrotic LV tissue showed minimal oxygen consumption comparable to media-only wells across all parameters, confirming that this assay reliably detects loss of mitochondrial function in non-viable tissue. In contrast, HOPE 20h tissue demonstrated robust basal and maximal OCR well above both control conditions, indicating preserved, metabolically active, viable myocardium despite extended preservation. This experiment was performed once (n=1 necrotic sample) as an internal proof-of-concept control. Data are presented as mean ± SEM.

**Supplemental Figure 3.**
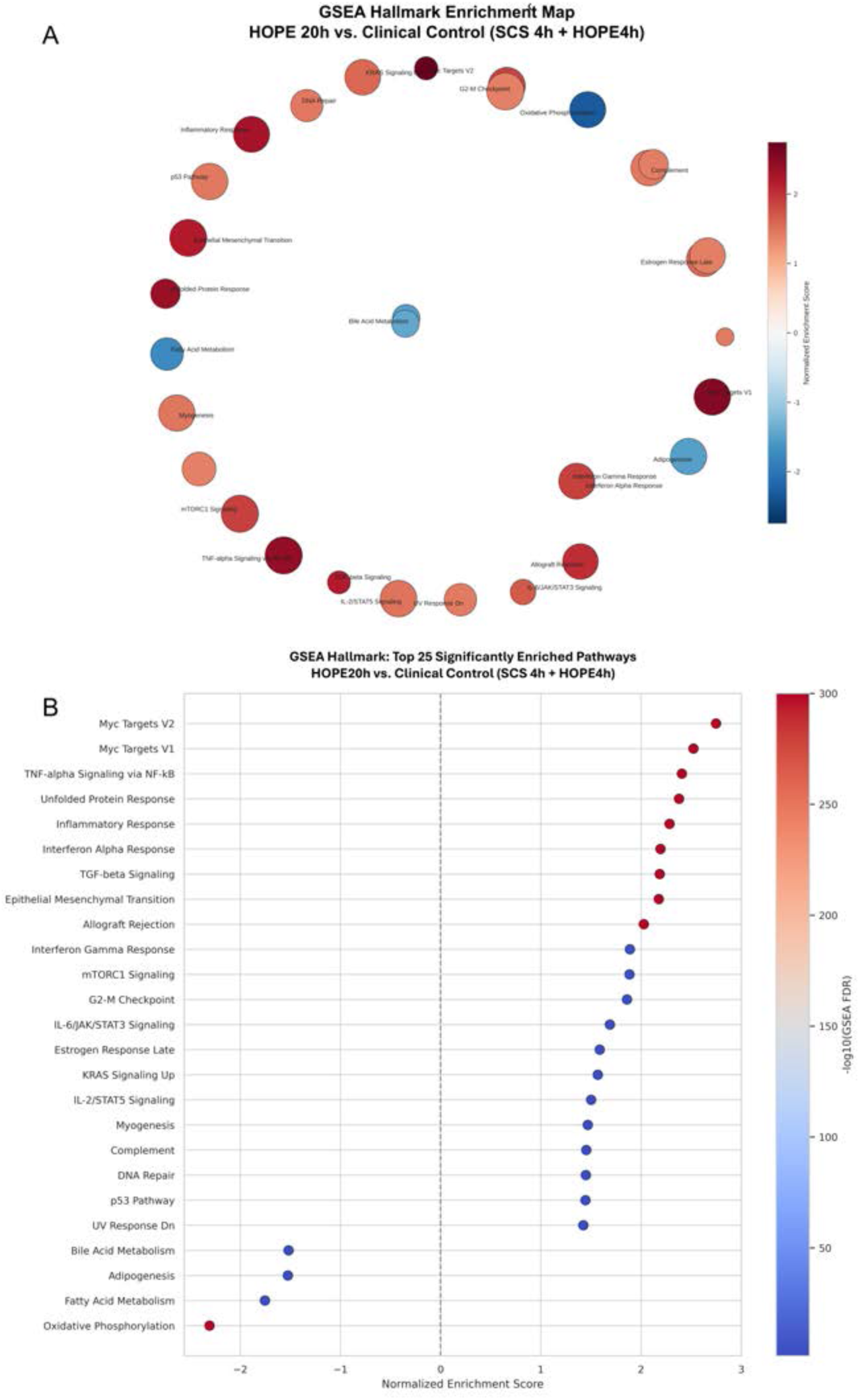
Transcriptomic GSEA Hallmark Enrichment. **(A)** Network visualization of enriched Molecular Signatures Database (MSigDB) Hallmark pathways. Each node represents a Hallmark gene set. **(B)** Ranked dot plot of top 25 significantly enriched Hallmark pathways according to NES.

**Supplemental Figure 4.**
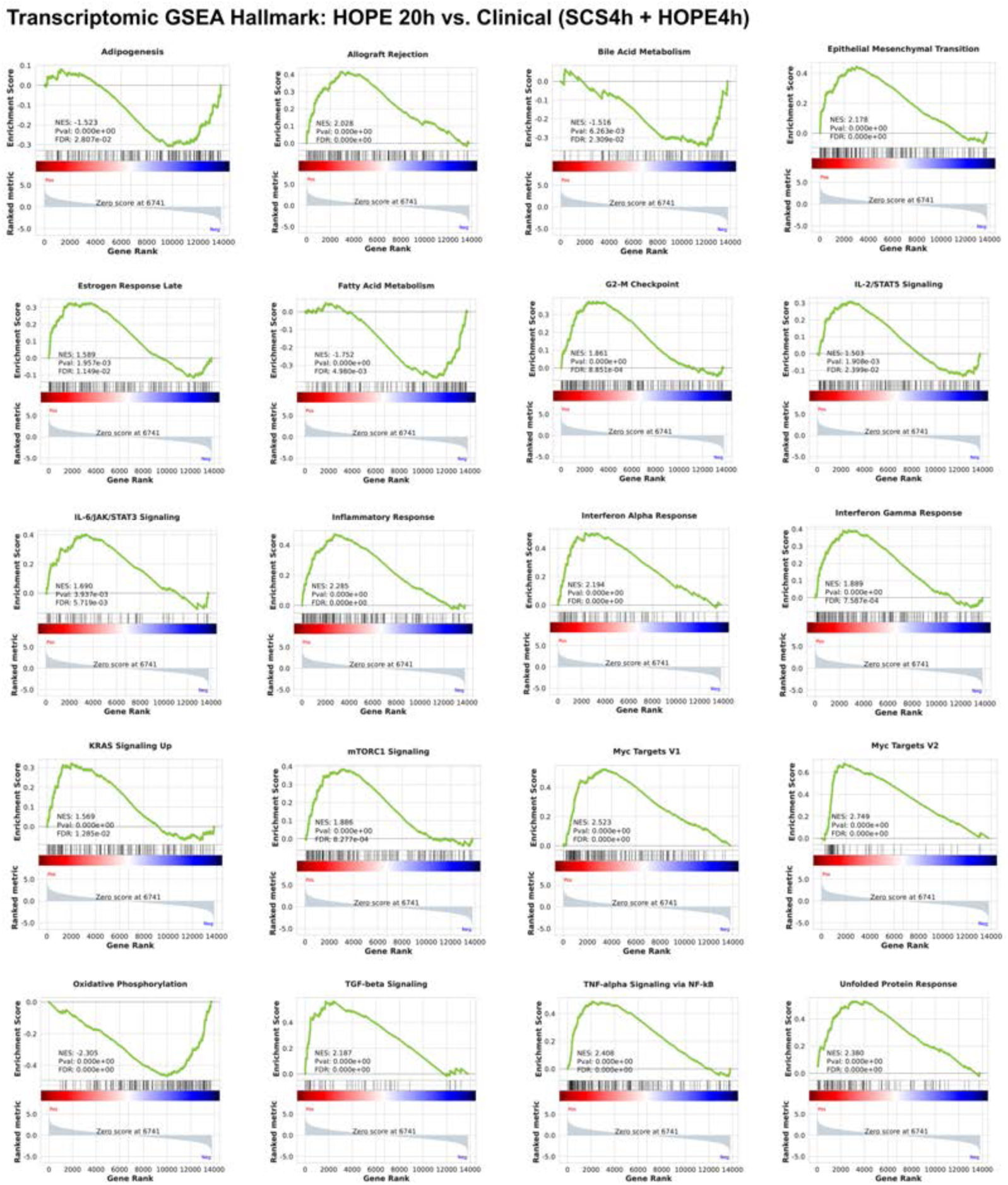
Top 20 Significantly Enriched Hallmark Gene Set Enrichment Analysis Reveals Distinct Inflammatory, Stress-Response, and Metabolic Transcriptional Signatures Following Extended Hypothermic Oxygenated Heart Preservation. Gene set enrichment analysis (GSEA) of bulk RNA sequencing data comparing extended HOPE 20 vs. Clinical Control (SCS4h + HOPE4h).

**Supplemental Figure 5.**
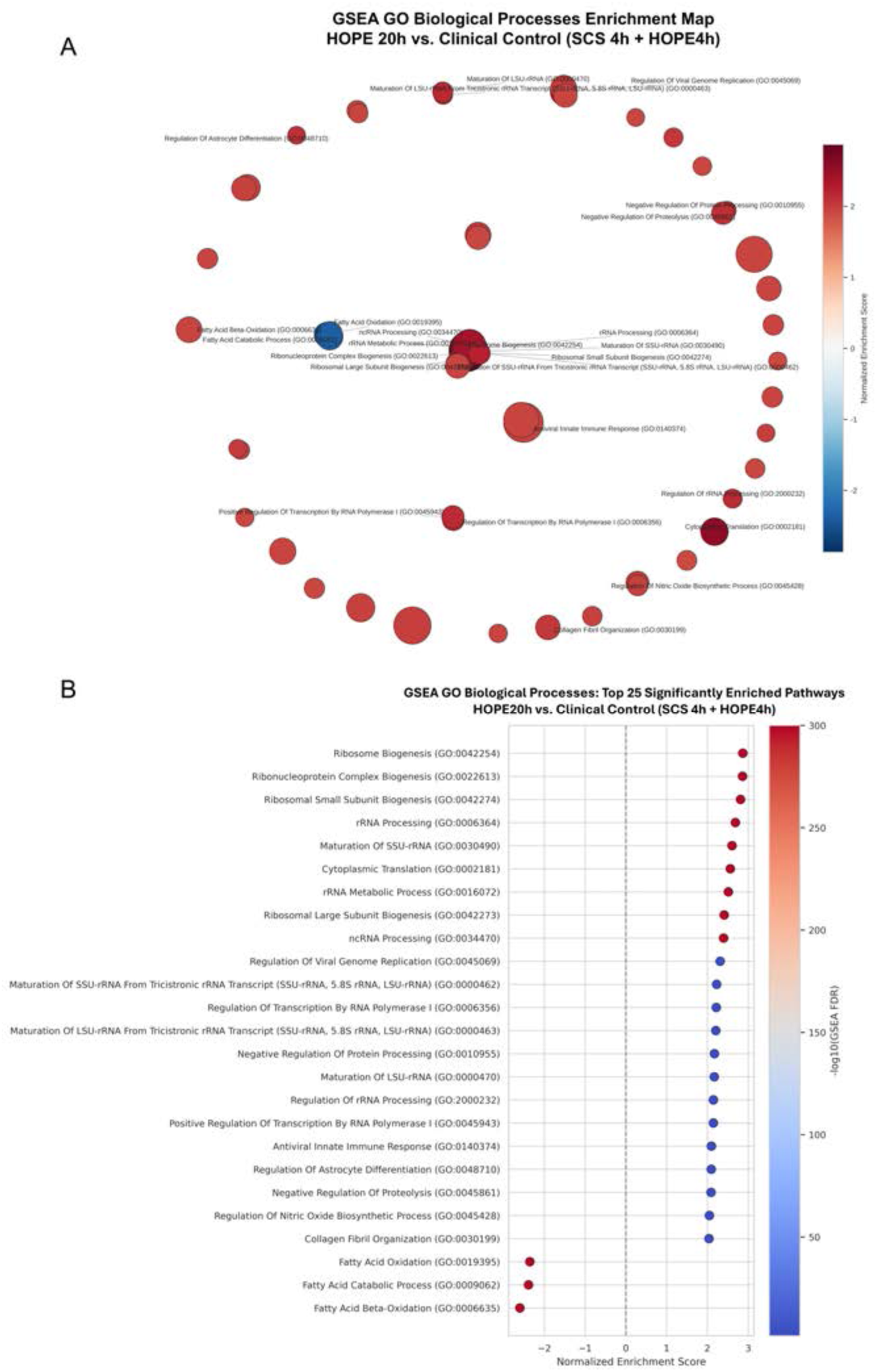
Transcriptomic GSEA Gene Ontology Biological Processes. **(A)** Enrichment map illustrating significantly enriched GO pathways. Each node represents an individual GO: Biological Process pathway, with node color corresponding to the normalized enrichment score (NES). **(B)** Ranked dot plot displaying the 25 significantly enriched GO pathways.

**Supplemental Figure 6.**
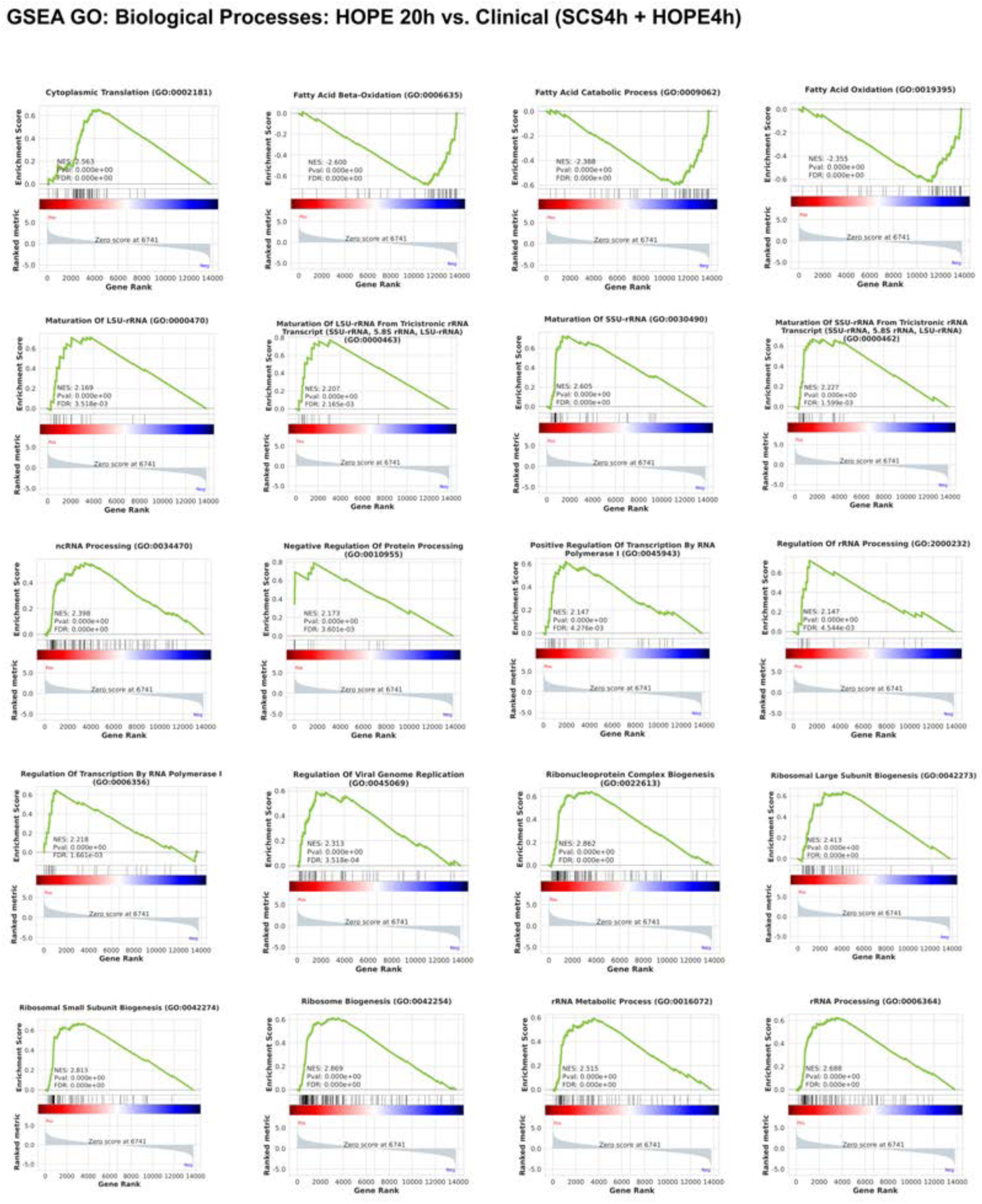
Top 20 Significantly Enriched Gene Ontology Biological Processes Analysis Identifies Enhanced Ribosomal Biogenesis and RNA Processing With Relative Depletion of Fatty Acid Metabolic Signatures. Gene set enrichment analysis (GSEA) of bulk RNA sequencing data comparing extended HOPE20h vs. Clinical Control (SCS4h + HOPE4h).

**Supplemental Figure 7.**
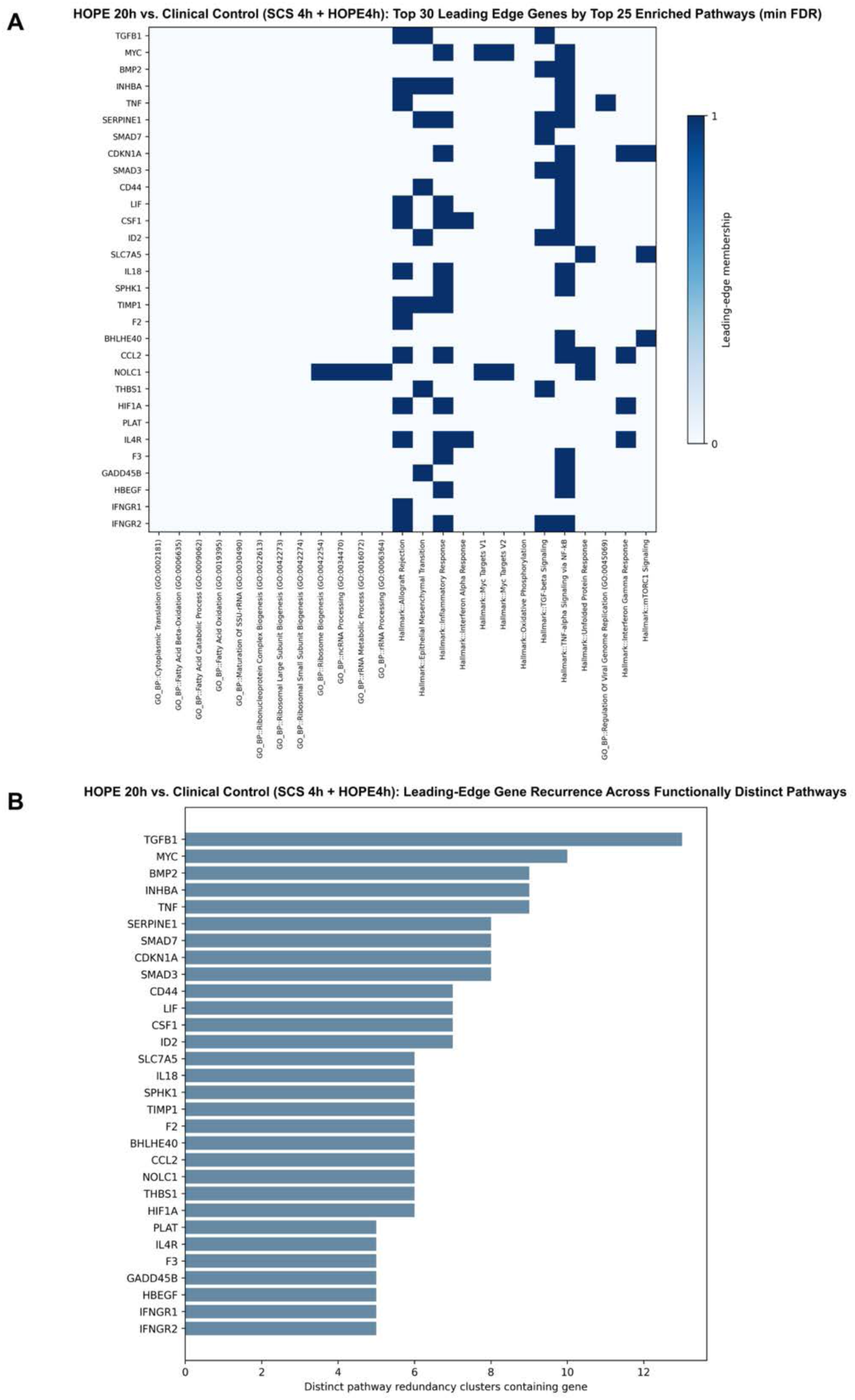
Redundancy-Aware Leading-Edge Gene Analysis Identifies Recurrent Transcriptional Contributors to Enriched Biological Pathways Following Extended HOPE. (A) Binary heatmap illustrating leading-edge membership of the 30 highest-ranked genes across the top 25 significantly enriched pathways ranked by minimum FDR. (B) Horizontal bar plot displaying the number of distinct pathway redundancy clusters containing each leading edge-gene.

**Supplemental Figure 8.**
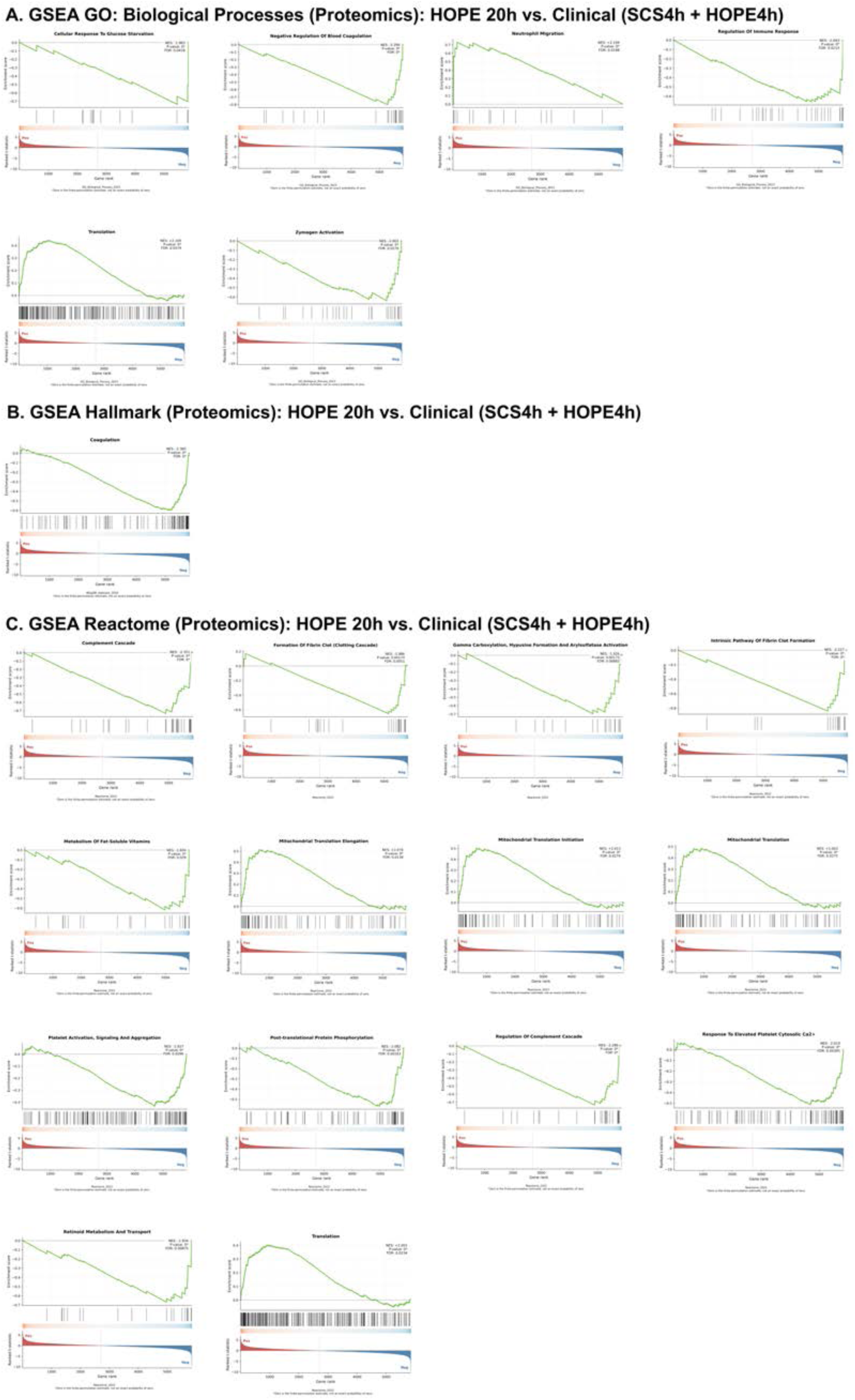
Gene Set Enrichment Analysis of Differential Protein Abundance Following Extended HOPE. (A) GO enrichment identified pathways (B) Hallmark enrichment (C) Reactome enrichment

**Supplemental Figure 9.**
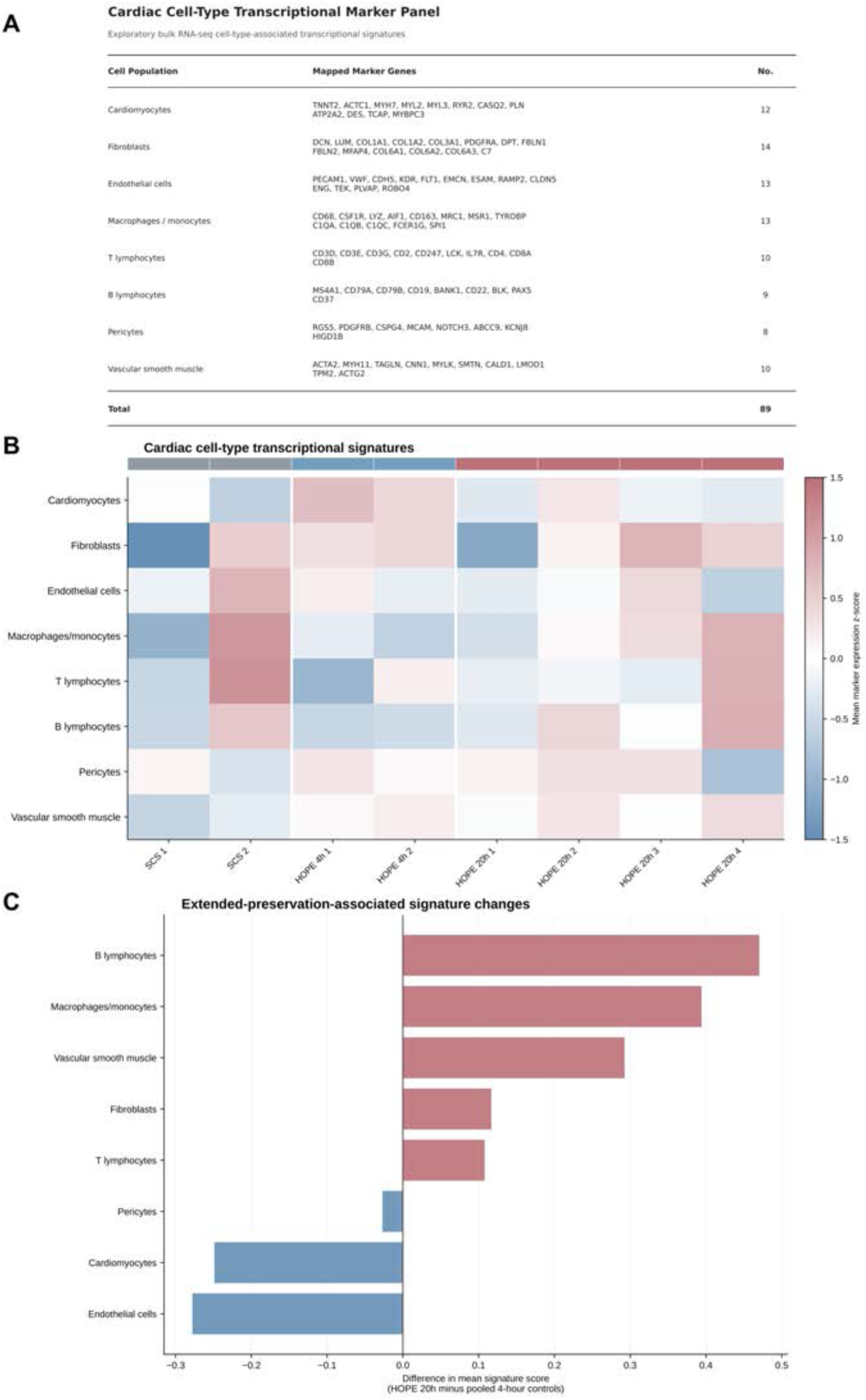
Cardiac Cell-Type-Associated Transcriptional Signatures Following Extended Hypothermic Oxygenated Heart Preservation. Exploratory cell-type-associated transcriptional signature analysis was performed using bulk RNA sequencing data comparing extended HOPE20h vs. Clinical Control (SCS4h + HOPE4h). (A) Summary of curated cell-type marker genes and their representation within the bulk RNA sequencing dataset. (B) Heatmap displaying cell-type-associated transcriptional signatures across individual preservation samples, including static cold storage (SCS4h; n=2), short-duration hypothermic oxygenated perfusion (HOPE4h; n=2), and extended hypothermic oxygenated perfusion (HOPE20h; n=4). (C) Differences in mean cell-type-associated transcriptional signature scores between HOPE20h and pooled clinical controls.

**Supplemental Table 1.**
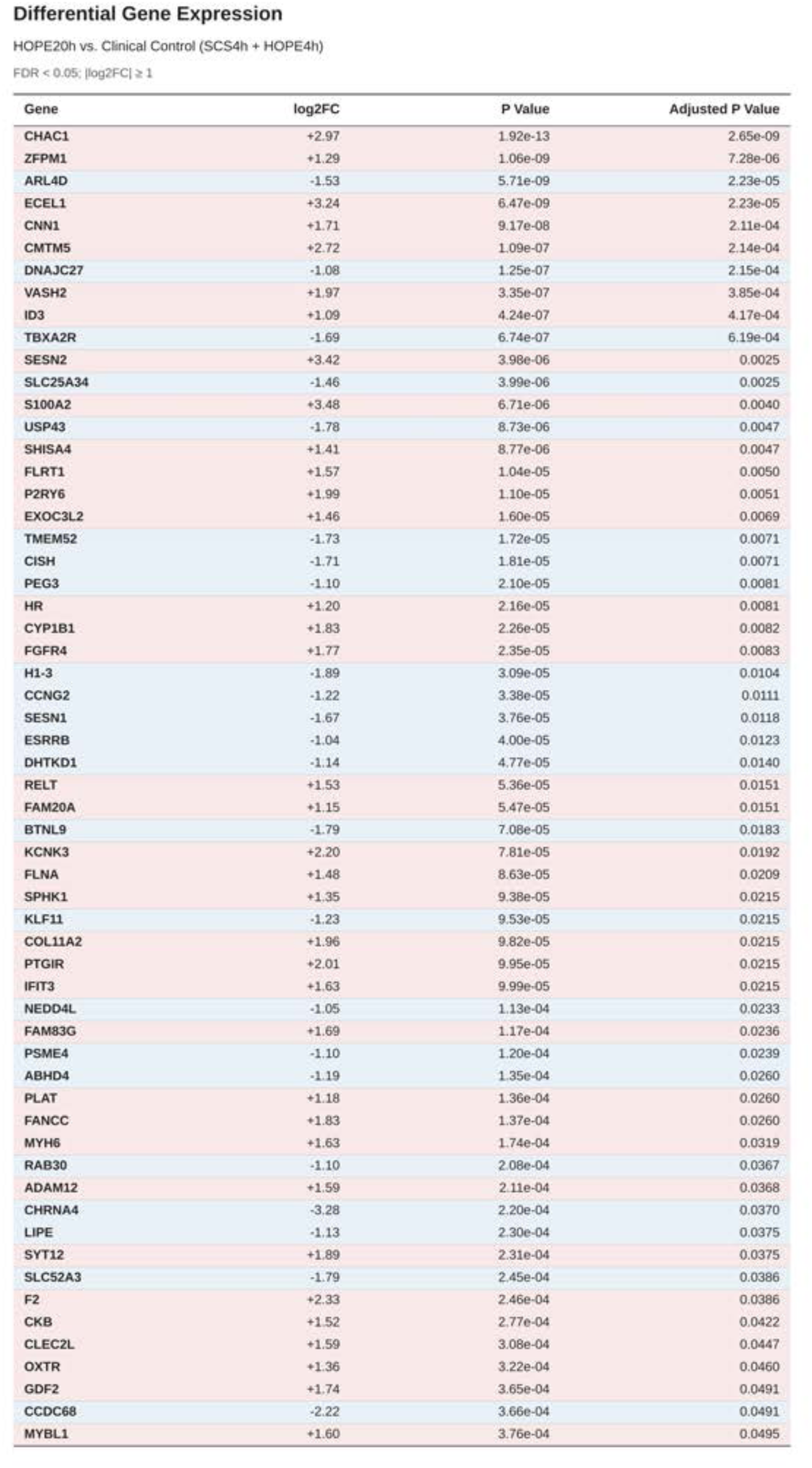
Differentially Expressed Genes. List of differentially expressed genes (DEG) with their respective log2FC, P-value and adjusted P-value. Red demonstrates upregulation in HOPE 20h, blue demonstrates down regulation in HOPE 20h.

| Gene | log2FC | P Value | Adjusted P Value |
|---|---|---|---|
| CHAC1 | +2.97 | 1.92e-13 | 2.65e-09 |
| ZFPM1 | +1.29 | 1.06e-09 | 7.28e-06 |
| ARL4D | -1.53 | 5.71e-09 | 2.23e-05 |
| ECEL1 | +3.24 | 6.47e-09 | 2.23e-05 |
| CNN1 | +1.71 | 9.17e-08 | 2.11e-04 |
| CMTM5 | +2.72 | 1.09e-07 | 2.14e-04 |
| DNAJC27 | -1.08 | 1.25e-07 | 2.15e-04 |
| VASH2 | +1.97 | 3.35e-07 | 3.85e-04 |
| ID3 | +1.09 | 4.24e-07 | 4.17e-04 |
| TBXA2R | -1.69 | 6.74e-07 | 6.19e-04 |
| SESN2 | +3.42 | 3.98e-06 | 0.0025 |
| SLC25A34 | -1.46 | 3.99e-06 | 0.0025 |
| S100A2 | +3.48 | 6.71e-06 | 0.0040 |
| USP43 | -1.78 | 8.73e-06 | 0.0047 |
| SHISA4 | +1.41 | 8.77e-06 | 0.0047 |
| FLRT1 | +1.57 | 1.04e-05 | 0.0050 |
| P2RY6 | +1.99 | 1.10e-05 | 0.0051 |
| EXOC3L2 | +1.46 | 1.60e-05 | 0.0069 |
| TMEM52 | -1.73 | 1.72e-05 | 0.0071 |
| CISH | -1.71 | 1.81e-05 | 0.0071 |
| PEG3 | -1.10 | 2.10e-05 | 0.0081 |
| HR | +1.20 | 2.16e-05 | 0.0081 |
| CYP1B1 | +1.83 | 2.26e-05 | 0.0082 |
| FGFR4 | +1.77 | 2.35e-05 | 0.0083 |
| H1-3 | -1.89 | 3.09e-05 | 0.0104 |
| CCNG2 | -1.22 | 3.38e-05 | 0.0111 |
| SESN1 | -1.67 | 3.76e-05 | 0.0118 |
| ESRRB | -1.04 | 4.00e-05 | 0.0123 |
| DHTKD1 | -1.14 | 4.77e-05 | 0.0140 |
| RELT | +1.53 | 5.36e-05 | 0.0151 |
| FAM20A | +1.15 | 5.47e-05 | 0.0151 |
| BTNL9 | -1.79 | 7.08e-05 | 0.0183 |
| KCNK3 | +2.20 | 7.81e-05 | 0.0192 |
| FLNA | +1.48 | 8.63e-05 | 0.0209 |
| SPHK1 | +1.35 | 9.38e-05 | 0.0215 |
| KLF11 | -1.23 | 9.53e-05 | 0.0215 |
| COL11A2 | +1.96 | 9.82e-05 | 0.0215 |
| PTGIR | +2.01 | 9.95e-05 | 0.0215 |
| IFIT3 | +1.63 | 9.99e-05 | 0.0215 |
| NEDD4L | -1.05 | 1.13e-04 | 0.0233 |
| FAM83G | +1.69 | 1.17e-04 | 0.0236 |
| PSME4 | -1.10 | 1.20e-04 | 0.0239 |
| ABHD4 | -1.19 | 1.35e-04 | 0.0260 |
| PLAT | +1.18 | 1.36e-04 | 0.0260 |
| FANCC | +1.83 | 1.37e-04 | 0.0260 |
| MYH6 | +1.63 | 1.74e-04 | 0.0319 |
| RAB30 | -1.10 | 2.08e-04 | 0.0367 |
| ADAM12 | +1.59 | 2.11e-04 | 0.0368 |
| CHRNA4 | -3.28 | 2.20e-04 | 0.0370 |
| LIPE | -1.13 | 2.30e-04 | 0.0375 |
| SYT12 | +1.89 | 2.31e-04 | 0.0375 |
| SLC52A3 | -1.79 | 2.45e-04 | 0.0386 |
| F2 | +2.33 | 2.46e-04 | 0.0386 |
| CKB | +1.52 | 2.77e-04 | 0.0422 |
| CLEC2L | +1.59 | 3.08e-04 | 0.0447 |
| OXTR | +1.36 | 3.22e-04 | 0.0460 |
| GDF2 | +1.74 | 3.65e-04 | 0.0491 |
| CCDC68 | -2.22 | 3.66e-04 | 0.0491 |
| MYBL1 | +1.60 | 3.76e-04 | 0.0495 |

**Supplemental Table 2.** Differential Protein Abundance. List of differential protein abundance following proteomic analysis with their respective log2FC, P-value and adjusted P-value. Red demonstrates increased abundance in HOPE 20h, blue demonstrates decreased abundance in HOPE 20h.

| **Gene / Protein** | **log2FC** | **P Value** | **Adjusted P Value** |
|---|---|---|---|
| **AMBP** | -1.10 | 1.41e-04 | 8.29e-09 |
| **MPO** | +1.26 | 2.23e-04 | 6.76e-08 |
| **PR39** | +1.02 | 3.96e-04 | 1.65e-06 |
| **LCN2** | +1.42 | 4.42e-04 | 2.49e-06 |
| **A0A287BNX9** | -3.02 | 4.95e-04 | 3.89e-06 |
| **ARAP1** | -2.39 | 0.0011 | 1.65e-04 |
| **S100A12** | +1.62 | 0.0014 | 2.91e-04 |
| **MRPL55** | +1.12 | 0.0014 | 3.08e-04 |
| **LOC100736951** | +2.11 | 0.0015 | 3.67e-04 |
| **HECTD3** | +1.61 | 0.0020 | 7.94e-04 |
| **PLAT** | -1.17 | 0.0022 | 0.0010 |
| **GPR107** | -1.46 | 0.0024 | 0.0013 |
| **SLA-7** | -4.60 | 0.0024 | 0.0014 |
| **A0A287AE25** | +1.39 | 0.0027 | 0.0017 |
| **CYBA** | +1.80 | 0.0032 | 0.0028 |
| **MMP9** | +1.34 | 0.0036 | 0.0036 |
| **RBBP6** | +1.54 | 0.0038 | 0.0042 |
| **IRAK4** | +1.39 | 0.0038 | 0.0043 |
| **S100A9** | +1.57 | 0.0045 | 0.0059 |
| **LTF** | +1.08 | 0.0046 | 0.0062 |
| **A0A5G2QBU3** | +1.69 | 0.0047 | 0.0063 |
| **DARS1.1** | +2.51 | 0.0050 | 0.0070 |
| **PTPN21** | +1.07 | 0.0050 | 0.0071 |
| **PMAP23** | +2.06 | 0.0052 | 0.0074 |
| **ATP5IF1** | +1.05 | 0.0052 | 0.0074 |
| **AZU1** | +1.12 | 0.0052 | 0.0075 |
| **BET1L** | +1.59 | 0.0053 | 0.0076 |
| **POMT2** | -1.69 | 0.0056 | 0.0084 |
| **ABHD18** | +3.12 | 0.0056 | 0.0085 |
| **RECQL5** | +3.00 | 0.0060 | 0.0102 |
| **SMG7** | +1.64 | 0.0071 | 0.0149 |
| **A0A8W4FGK7** | -1.04 | 0.0075 | 0.0168 |
| **CD177** | +1.82 | 0.0082 | 0.0199 |
| **A0A8W4FEL9** | -1.13 | 0.0084 | 0.0210 |
| **A0A287B1P1** | +1.01 | 0.0085 | 0.0214 |
| **MBP** | +2.43 | 0.0087 | 0.0224 |
| **ALOX15** | +1.49 | 0.0097 | 0.0280 |
| **SOS1** | +3.49 | 0.0108 | 0.0348 |
| **MYL1** | -1.76 | 0.0109 | 0.0355 |

**Supplemental Table 3.** Integrated Comparison of Transcriptomic and Proteomic Expression Following Extended Hypothermic Oxygenated Heart Preservation. 50 highest-ranking by minimum FDR matched RNA-protein pairs demonstrating FDR <0.05 in at least one omics layer.

| Gene | RNA log2FC | RNA FDR | Protein log2FC | Protein FDR |
|---|---|---|---|---|
| **USP46** | -0.09 | 0.928 | **-0.96** | **<0.001** |
| **LCN2** | +0.36 | 0.759 | **+1.42** | **<0.001** |
| **UBE3B** | -0.12 | 0.859 | **-0.81** | **<0.001** |
| **ARAP1** | +0.17 | 0.818 | **-2.39** | **<0.001** |
| **CNN1** | **+1.71** | **<0.001** | +0.37 | 0.919 |
| **LRRC2** | **-1.00** | **<0.001** | +0.21 | 0.933 |
| **C7** | +0.30 | 0.841 | **-0.88** | **<0.001** |
| **RPL37** | +0.16 | 0.837 | **+0.79** | **<0.001** |
| **PLAT** | **+1.17** | **0.026** | **-1.17** | **0.0010** |
| **SLA-7** | -0.23 | 0.782 | **-4.60** | **0.0014** |
| **CYBA** | +0.26 | 0.773 | **+1.80** | **0.0028** |
| **APOE** | +1.04 | 0.196 | **-0.82** | **0.0031** |
| **PTGS1** | -0.22 | 0.794 | **+0.86** | **0.0034** |
| **MARCKS** | +0.04 | 0.974 | **-0.55** | **0.0036** |
| **MMP9** | +0.61 | 0.640 | **+1.34** | **0.0036** |
| **NAMPT** | **-0.79** | **0.0037** | -0.10 | 0.897 |
| **RBBP6** | -0.01 | 0.987 | **+1.54** | **0.0042** |
| **IRAK4** | +0.29 | 0.680 | **+1.39** | **0.0043** |
| **PRTN3** | +0.68 | — | **+0.82** | **0.0044** |
| **ANTKMT** | -0.10 | 0.837 | **-0.92** | **0.0045** |
| **LRRK2** | -0.06 | 0.953 | **+0.74** | **0.0047** |
| **AARS1** | **+0.69** | **0.0051** | -0.17 | 0.884 |
| **STAB1** | -0.17 | 0.765 | **+0.50** | **0.0056** |
| **MASP1** | **-0.77** | **0.0061** | -0.12 | 0.923 |
| **LTF** | -0.01 | 0.995 | **+1.08** | **0.0062** |
| **PLIN3** | -0.84 | 0.087 | **-0.53** | **0.0062** |
| **PLIN2** | -0.55 | 0.491 | **-0.82** | **0.0072** |
| **H1-3** | **-1.89** | **0.0072** | -0.01 | 0.942 |
| **QSOX1** | -0.58 | 0.270 | **-0.88** | **0.0075** |
| **BET1L** | +0.07 | 0.909 | **+1.59** | **0.0076** |
| **TXNIP** | -0.92 | 0.318 | **-1.00** | **0.0078** |
| **SELENOK** | -0.07 | 0.926 | **+0.68** | **0.0083** |
| **POMT2** | -0.12 | 0.909 | **-1.69** | **0.0084** |
| **SESN1** | **-1.67** | **0.0097** | -0.08 | 0.926 |
| **EDNRA** | -0.66 | 0.302 | **-0.94** | **0.012** |
| **ESRRB** | **-1.04** | **0.013** | -0.85 | 0.353 |
| **ALG11** | +0.19 | 0.782 | **+0.50** | **0.013** |
| **C2** | +0.04 | 0.977 | **-0.89** | **0.014** |
| **BTNL9** | **-1.79** | **0.015** | -2.05 | 0.066 |
| **SMG7** | +0.05 | 0.920 | **+1.64** | **0.015** |
| **IKBKB** | +0.15 | 0.791 | **-0.52** | **0.015** |
| **SMYD5** | +0.03 | 0.965 | **-0.51** | **0.017** |
| **HEMK1** | -0.26 | 0.660 | **-0.84** | **0.017** |
| **HYAL2** | **-0.77** | **0.019** | -0.25 | 0.870 |
| **PATL1** | -0.06 | 0.911 | **-0.70** | **0.019** |
| **FLNA** | **+1.48** | **0.019** | -0.02 | 0.944 |
| **IFIT3** | **+1.63** | **0.019** | -0.28 | 0.664 |
| **POSTN** | -1.10 | 0.132 | **-0.88** | **0.021** |
| **MBP** | +0.34 | 0.773 | **+2.43** | **0.022** |
| **PSME4** | **-1.10** | **0.024** | -0.03 | 0.934 |

## Supplemental Materials and Methods

Our in vivo large animal model of HT was carried out using female 4 month-old Yorkshire pigs weighing approximately 60-65kg. All hearts were harvested using a standard, clinically accepted procurement technique with bicaval implantation following donation after brain death (DBD). Hearts were stored in SCS for 4 hours or HOPE for 4 or 20 hours before implant. Due to institutional policy, donor pigs were not subject to any procedure to inflict brain death. The protocol was approved by Columbia’s Institute for Comparative Medicine (ICM) under protocol number AC-AACA3703 and all procedures were performed at Columbia University Medical Center.

### Preoperative Donor and Recipient Preparation

Prior to transplant procedure, three Yorkshire pigs 60-70kg were sedated and 5mL of blood is withdrawn for blood typing. Eldon Blood Typing Kit (Eldon Biologicals Home Kit 2511) was used to determine blood type for donor-recipient matching. All animals were made NPO at least 8 hours prior to surgery. Both the donor and recipient pig were sedated using 4.5 mg/kg of Telazol and 1.0 mg/kg of Propofol with maintenance by administering 1%–5% inhaled isoflurane. Pigs were orally intubated and mechanically ventilated with a tidal volume of 10 mL/kg/min, a rate of 10-15 breaths per minute was maintained for the remainder of the procedure. A water-based veterinary ointment was applied directly to the pig’s corneas for lubrication. IV catheters were placed within the bilateral auricular veins. Muscle paralysis of the donor was achieved using 0.2 mg/kg of vecuronium. Fentanyl infusion at 25-100 μg/h given for analgesia. Monitoring equipment for electrocardiogram monitoring, non-invasive blood pressure, and pulse oximetry were placed. Mechanical ventilation with a tidal volume of 10 mL/kg/min, a rate of 10–15 breaths per minute was maintained for the remainder of the procedure. Heart rate (60-100 bpm) and systolic blood pressure (90-130 mmHg) were maintained. Pigs were positioned supine, scrubbed with a Providine-iodine 8.3% with isopropyl alcohol 72.5% antiseptic solution, and draped for median sternotomy in standard fashion.

### Donor Cardiectomy

Once an appropriate plane of anesthesia was reached, a median sternotomy was performed. A sternal retractor was placed, the thymus was excised, and a pericardial well was created with traction stitches. A baseline functional assessment of the donor heart was performed. Echocardiography, using long and short axis views from the epicardial surface, was used to assess baseline function. Once preliminary assessment was obtained, the heart was prepared for procurement. Heparin was administered (350 units/kg). The right atrium is cannulated with a dual-stage 30-36Fr venous cannula, clamped, and connected to a cell saver. The proximal ascending aorta is cannulated with a 9 Fr antegrade cardioplegia cannula. After sufficient cardiac and great vessel dissection, the right atrial cannula was opened. The left atrial appendage was also opened widely; the ascending aorta was cross-clamped, and cold cardioplegia was delivered. Cold saline slush was used topically to cool the heart once an arrest was confirmed. After monitoring for LV distention and adequate root pressure throughout the period of cardioplegia delivery, the heart was excised in standard fashion. Care was taken to harvest sufficient aortic length to facilitate loading into the XVIVO Heart Assist Transport. Prior to initiating HOPE, the cardioplegia cannulation site was closed and reinforced, the left atrial appendage was closed with running suture, a 5F sheath was secured within the appendage suture line for placement of the solid-state catheter, and a silastic shunt is placed across the mitral valve and sutured in place to ensure adequate LV venting.

### Cardioplegic Conditions

2 liters of histidine-tryptophan-ketoglutarate (HTK) was used for cardioplegic arrest for hearts allocated to the SCS group. SCS hearts were submerged in HTK solution throughout preservation and stored at 4-6 °C. The hearts allocated to the HOPE group were arrested using XVIVO Heart Solution. XVIVO Heart Solution was also used to prime the perfusion box with 350cc of packed RBCs. One of the 1000cc priming bags of XVIVO Heart Solution was supplemented with 3ml of XVIVO Heart Solution Supplement, 20 IU Insulin, 50mg imipenem, 5000 IU unfractionated heparin and 10mmol of potassium. 100mEq of sodium bicarbonate is added to preservation box. HOPE hearts stored for 4 hours were arrested with 1L XHS. HOPE hearts stored for 20 hours were arrested with 2L XHS. Preservations were set at standard XVIVO recommendations: aortic root pressure of 20 mmHg and temperature of 8 °C.

### Recipient Implant

Recipient pigs were anesthetized and prepped in similar fashion to the donor as described above. Femoral vein central line insertion and arterial catheterization was performed using ultrasound guidance and Seldinger’s technique. All inotropes and vasopressors were administered centrally.

Urethral catheterization in female swine is challenging due to anatomical conformation. Suprapubic access provides a reliable and simple alternative for direct bladder cannulation and Foley placement prior to recipient operation^30^. A 5cm inferior laparotomy was made, and the bladder was eviscerated. A catheter is inserted directly to the inferior, dependent portion of the bladder. The skin was closed and the catheter was secured to the skin.

Median sternotomy was performed in standard fashion, the thymus was removed, and a pericardial well is created. Heparin was given with confirmation of ACT > 350. Aortic and bicaval cannulas for CPB were inserted, and CPB was initiated. The distal ascending aorta was cross clamped, and recipient cardiectomy was performed in standard fashion. The left hemi-azygous vein was ligated.

The donor heart was removed from SCS or HOPE preservation and brought onto the field. Standard bicaval implantation was performed. Prior to completion of the left atrial anastomosis, a solid-state pressure catheter was inserted through the previously placed sheath in the left atrial appendage into the left atrium and advanced beyond the mitral valve orifice into the left ventricle.

Following completion of the final anastomosis and removal of the aortic cross-clamp, the heart was reperfused while pigs remained on cardiopulmonary bypass. Defibrillation was required to restore sinus rhythm in most cases, typically necessitating 15–30 minutes of intermittent defibrillation attempts with 10-30J using internal paddles. During the reperfusion period; spontaneous return of sinus rhythm without defibrillation occurred in one 20-hour HOPE experiment. Animals were maintained on full cardiopulmonary bypass support for one hour following reperfusion before initiating a slow wean, with flow reduced in a stepwise fashion at approximately 500-1000 mL/min decrements. Inotropic and vasopressor support was titrated as needed during weaning as described above. In our initial two experiments, pigs were cooled to 34°C during bypass; in later experiments, active cooling was discontinued to reduce shivering, which seemed to limit lactatemia once bypass was initiated. Vecuronium was administered for neuromuscular blockade to further suppress shivering once CPB was initiated.

The chest was left open throughout the experiment. Animals were monitored for a 3-hour terminal observation period. Vitals were recorded at 15-minute intervals throughout the monitoring period. Concurrent infusion rates of epinephrine, dobutamine, vasopressin, norepinephrine, amiodarone, and any additional vasoactive infusions were recorded at each time point. Right atrial pressure, urine output, arterial blood gas, and serum chemistries were assessed at hourly intervals. Short axis epicardial echocardiography was performed hourly to assess graft function. Total ‘ischemic time’ was calculated as the interval between donor aortic cross-clamp application and removal of the recipient aortic cross-clamp.

### Veterinary Intraoperative Management

*Anesthetic Maintenance:* Animals were maintained on a continuous rate infusion (CRI) of fentanyl and propofol with low-concentration isoflurane 2-5%. Ketamine CRI was sometimes added, particularly when hemodynamic support was needed while minimizing isoflurane and opioid requirements. During induced hypothermia on cardiopulmonary bypass, CRI rates was reduced proportionally to account for decreased metabolic demand.

#### Immunosuppression

Methylprednisolone 1 g by IV was administered as a bolus at the start of the procedure. If mycophenolate mofetil was indicated per protocol, it was administered via orogastric gavage using a large-bore rubber catheter.

#### Hemodynamic Support

Epinephrine and norepinephrine were prepared in advance as the primary vasopressors. Initial management of hypotension consisted of reducing isoflurane while increasing propofol CRI and optimizing volume status. Vasopressors were titrated if hypotension persisted or if myocardial contractility was impaired.

#### Anticoagulation and Reversal

Heparin was administered immediately prior to cannulation for cardiopulmonary bypass, with activated clotting time (ACT) monitored by the perfusion team targeting 350+ seconds. Additional heparin boluses were given as needed to maintain therapeutic anticoagulation. Upon separation from bypass, protamine was dosed based on the total cumulative heparin administered and infused slowly to minimize adverse hemodynamic effects.

#### Temperature Management

Intentional hypothermia was induced by the perfusion team during bypass, with core body temperature permitted to decrease to approximately 34°C (94°F). Active warming devices were discontinued upon initiation of bypass and restarted during the rewarming phase prior to separation. Temperature decline during this period was expected and required no intervention. In latter experiments, cooling by CPB was terminated, lactate elevation was not observed.

#### Antiarrhythmic Therapy

Lidocaine IV bolus (1mg/kg) was given for acute arrhythmia management. Amiodarone was used for antiarrhythmic prophylaxis. It must be diluted to 2 mg/mL in 5% dextrose in sterile water. A loading dose of 2 mg/kg is infused over approximately 20 minutes, followed by a maintenance infusion of 1 mg/kg/hr.

#### Neuromuscular Blockade

Vecuronium was administered when neuromuscular blockade was desired, particularly to suppress shivering during hypothermia. A CRI of 4–8 mL/hr was typical, with supplemental boluses of 2–6 mL given for breakthrough muscular activity after confirming adequate anesthetic depth. When the animal is paralyzed and fully supported on bypass, anesthetic depth was confirmed with firm pressure applied between the digits of a forelimb.

#### Ventilator Management

Mechanical ventilation was maintained throughout the procedure, including during cardiopulmonary bypass, with positive end-expiratory pressure (PEEP) set at 4 cmH₂O. Tidal volume and respiratory rate was reduced during bypass to minimize barotrauma while preventing atelectasis.

### Clinical Endpoints Methods

Cardiac graft function was serially assessed using echocardiography, invasive hemodynamic monitoring, and a composite vasoactive support score. Visual ejection fraction (EF), fractional area change, and left ventricular end diastolic diameter (LVEDD) were measured by echocardiography at donor baseline (prior to cardiectomy) and at 0.5, 1.5, and 2.5 hours following separation from cardiopulmonary bypass by parasternal short axis, via direct epicardial contact. Left ventricular contractility was assessed via the left ventricular solid-state pressure catheter at 1 and 2 hours off bypass. Central venous pressure and systolic blood pressure were recorded at 1 and 2 hours following separation from bypass. A combined drip score, reflecting cumulative vasoactive and inotropic support, was calculated at 1 and 2 hours off bypass by adding epinephrine and levophed dosing. Statistical comparisons between groups at each timepoint were performed using ANOVA. Hemodynamics

### Optical Flow: Quantified Epicardial Motion

High-definition video of cardiac contraction was obtained at baseline, prior to donor cardiectomy, and after implant throughout the monitoring period (1.5h and 2.5h off CPB). Once obtained, the videos were stabilized and analyzed using a custom Python pipeline to quantify epicardial motion and estimate contractile function^29^. Videos were converted to grayscale, and frame-frame pixel displacement in a standardized area at interventricular groove that includes both the right ventricle (RV) and the left ventricle (LV) was estimated using the Farnebäck dense optical flow algorithm in OpenCV^30^.

Motion magnitude was averaged and converted from pixels/frame to mm/s using frame rate metadata and a calibration factor in the field of view. Raw velocity was smoothed through a Savitzy-Golay filter to obtain average velocity magnitudes throughout the cardiac cycle. To isolate systolic function, velocity values during each ventricular contraction were plotted, and the peak velocities were identified and averaged to obtain mean peak velocity (MPV) of each group. Furthermore, velocity plots were mathematically differentiated to obtain mean peak acceleration (MPA) of the epicardium during contraction. Statistical significance was calculated with one-way ANOVA.

### Tissue Collection

At the conclusion of the three-hour post-wean monitoring period, blood and tissue collection was performed immediately. Hearts were explanted while still warm and beating and immediately processed for downstream analyses, including flow cytometry, histology, mitochondrial respirometry (Seahorse assay) and RNA isolation and sequencing.

The explanted heart was systematically biopsied using a 8mm biopsy punch across left ventricular (LV) free wall and interventricular septum (IVS).

### Histological Analysis

Myocardial tissue obtained from LV and IVS regions was fixed in 10% formalin (4% formaldehyde) for < 24 hours, then transferred to 70% ethanol and embedded in paraffin. Next, tissue blocks were sliced using a microtome and 10 µm-tick sections were stained with Hematoxylin and Eosin (H&E) and Masson’s trichrome for histological evaluation. Additional 5 µm-tick blank slices were generated for chromogenic terminal deoxynucleotidyl transferase dUTP nick-end labeling (TUNEL) with HRP-DAB system. Histological specimen preparation and staining were performed in the Molecular Pathology Shared Resource of the Herbert Irving Comprehensive Cancer Center at Columbia University.

### Flow cytometry

Myocardial biopsies of 100 mg were collected from the IVS region, washed immediately in PBS 1x (Corning) and minced into 2 mm^3^ using a scalpel blade in sterile conditions under the biosafety cabinet. Tissues were then transferred into 15 ml tubes containing a warm enzymatic solution consisting of DMEM Base (Gibco, 10313-021) with 450 U/ml Collagenase I (Worthington, LS004196), 60 U/ml DNase I (Millipore, DN25) and 60 U/ml hyaluronidase (Worthington, LS002592) and incubated in a rotating mixer in agitation at 65/70 rpm at 37 °C for 1 hour. The digestion was stopped by the addition of 8 ml of HBB buffer, consisting of 2% FBS (Sigma-Aldrich, 12306C) and 0.2% BSA (Prometheus, 25-529) in Hanks′ Balanced Salt solution (Gibco, 14175-095), and the single cells suspension was filtered using a 100 µm cell strainer to filter out undigested material and centrifuged at 350g for 5 min at 4 °C. Next, the pellet was resuspended in ACK lysing buffer (Gibco, A10492) to lyse red blood cells, incubated for 5 minutes at room temperature, washed with 9 ml of DMEM Base and centrifuged at 350g for 5 min at 4 °C. After one wash with PBS 1x, cells were incubated for 10 minutes at 4°C in 1 ml of Blocking Solution, consisting of 0.5% BSA in PBS. Then, 2 ml of PBS 1x were added to the tube and the cellular solution was split equally into three FACS tubes. Cells were centrifuged at 500g for 5 min at 4°C and the pellet from each FACS tube was resuspended in either 300 µl of Calcein AM (1:500) (BD Pharmingen, 564061) and DRAQ5 (1:2000) (Biolegend, 424101) in PBS, or only DRAQ5 (1:2000) in PBS (Calcein-negative control), or PBS only (unstained control). Samples were incubated for 30 minutes protected from light at 4°C, then washed with PBS 1x, centrifuged at 500g for 5 min at 4°C and finally resuspended in 300 µl of PBS. Cells were analyzed using the NovoCyte Penteon Flow Cytometer Systems 5 Lasers (Agilent) and data analysis was performed with the NovoExpress flow cytometry software. First, single particles were isolated from duplets by plotting Forward Scatter Area (FSC-A) against Forward Scatter Height (FSC-H) and gating only the events that localized along the diagonal line. Next, nucleated events were identified by DRAQ5 positivity, and, among them, metabolically active cells were identified by Calcein-AM positive staining. Calcein-AM is a cell-permeant probe that emits a fluorescent signal upon cleavage by cellular esterases and is retained within cells with intact membranes. Calcein-AM signal is proportional to cell viability and metabolic activity, and it normally decreases in low viable cell samples.^31^ Unstained and Calcein-AM negative controls have been used to exclude false positive background signals.

### Extracellular Flux (Seahorse) Mitochondrial Respirometry

#### Precision-Cut Myocardial Slice Preparation

Left ventricular (LV) free wall biopsies were obtained using a full thickness 8 mm punch and washed in 1× PBS. Biopsy cores were embedded in 3% (w/v) low-gelling-temperature agarose prepared in Tyrode’s solution and mounted on a vibrating microtome fitted with a metal blade. Tissue was sectioned at 150 µm thickness using an advance speed of 1 and oscillation setting of 6^24,30^. Slices were collected and maintained in cardioplegia solution on ice until further processing. Individual precision-cut slices were further trimmed to 3 mm diameter mini-slices using a biopsy punch, transferred to a wash buffer containing 2% P/S, 5ug/ml gentamycin, 1.25 ug/mL amphotericin B in Advanced DMEM and washed twice for 5 minutes each. Finally, slices were incubated overnight in Advanced DMEM containing 10% FBS, 1% L-glutamine, 2% P/S, 5 ug/ml gentamycin, 1.25 ug/mL amphotericin B.

#### Seahorse Microplate Preparation

Agilent Seahorse XFe96 sensor cartridges were hydrated with 200 µL of tissue-culture-grade deionized water per well and incubated overnight in a non-CO₂ incubator at 37°C. On the day of the assay, hydration water was replaced with 200 µL of XF Calibrant Solution per well. XF96 cell culture microplates were coated with Corning Cell-Tak tissue adhesive (3.75 µg per well, diluted in deionized water with 0.1 M sodium bicarbonate) for a minimum of 20 minutes to achieve a surface coverage of 5 µg/cm², then washed and allowed to dry prior to slice loading. nIndividual 3 mm × 150 µm myocardial mini-slices were transferred into Cell-Tak-coated wells containing 20 µL PBS to maintain slice flatness. PBS was removed, and slices were allowed to adhere to the well surface for 1–2 minutes. Warmed complete Seahorse XF DMEM assay medium (supplemented with 10 mM glucose, 1 mM pyruvate, and 2 mM L-glutamine) was then added to each well to a final volume of 180 µL, taking care to avoid slice detachment.^31^ Four wells per plate containing assay medium without tissue served as background. The loaded microplate was incubated for 45–60 minutes in a non-CO₂ incubator at 37°C prior to the assay to allow temperature and pH equilibration.

#### Mito Stress Test

Mitochondrial function was assessed using a modified Seahorse XF Cell Mito Stress Test (Agilent) on an XFe96/XF Pro Analyzer. Oligomycin was omitted from the standard protocol, as precision-cut myocardial slices have been reported not to respond reliably to oligomycin dosing ^32,33^. Carbonyl cyanide-4 (trifluoromethoxy)phenylhydrazone (FCCP, Port A) and a combination of rotenone and antimycin A (Rot/AA, Port B) were sequentially injected. FCCP was loaded to a final well concentration of 5 µM and Rot/AA to a final well concentration of 2 µM.^24^ The sensor cartridge was calibrated for 15 minutes prior to loading the tissue culture microplate. Oxygen consumption rate (OCR) and extracellular acidification rate (ECAR) were measured over 8 mix–wait–measure cycles (3 minutes mix, 2 minutes wait, 3 minutes measure per cycle)^32,24^. Basal OCR and ECAR were recorded prior to FCCP injection; maximal OCR and ECAR were recorded following FCCP-induced uncoupling; and non-mitochondrial OCR was recorded following Rot/AA administration. OCR and ECAR values were normalized to individual slice volume, calculated from slice diameter (3 mm) and thickness (150 µm). Raw data were exported and analyzed using Seahorse Analytics software, Microsoft Excel, and GraphPad Prism.

### RNA Extraction and Bulk RNA Sequencing

IVS tissue biopsies of approximately 200 mg were homogenized in 1 mL of TRIzol reagent (Invitrogen, Cat. N. 15596026) and stored at −80°C until RNA extraction.

#### Extraction

RNA extraction and sequencing were performed by GENEWIZ® NGS Services from Azenta Life Sciences (South Plainfield, NJ, USA).

Total RNA was extracted using Qiagen Rneasy Plus Universal Mini kit (Qiagen, Hilden, Germany) following manufacturer’s instructions.

#### Library Preparation

RNA samples were quantified using Qubit 4.0 Fluorometer (ThermoFisher Scientific, Waltham, MA, USA) and RNA integrity was checked using TapeStation (Agilent Technologies, Palo Alto, CA, USA). ERCC RNA Spike-In Mix (Cat: #4456740) from ThermoFisher Scientific was added to normalized total RNA prior to library preparation following manufacturer’s protocol.

RNA sequencing libraries were prepared using the NEBNext Ultra II RNA Library Prep Kit for Illumina and NEBNext Poly(A) mRNA Magnetic Isolation Module (New England Biolabs, Ipswich, MA, USA), according to manufacturer’s recommendations. Briefly, mRNAs were initially enriched with Oligod(T) beads. Enriched mRNAs were fragmented for 15 minutes at 94 °C. First strand and second strand cDNA were subsequently synthesized. cDNA fragments were end-repaired and adenylated at 3’ends, and universal adapters were ligated to cDNA fragments, followed by index addition and library enrichment by PCR with limited cycles. The sequencing library was validated on the Agilent TapeStation, and quantified by using Qubit 4.0 Fluorometer as well as by quantitative PCR (KAPA Biosystems, Wilmington, MA, USA).

#### Sequencing

The sequencing libraries were multiplexed and clustered onto a flowcell. After clustering, the flowcell was loaded on the Illumina NovaSeq instrument according to manufacturer’s instructions. The samples were sequenced using a 2×150bp Paired End (PE) configuration. Image analysis and base calling were conducted by the NovaSeq Control Software. Raw sequence data (.bcl files) generated from Illumina NovaSeq was converted into fastq files and de-multiplexed using Illumina bcl2fastq 2.20 software. One mismatch was allowed for index sequence identification.

#### Data Analysis

Differential gene expression analysis was performed using PyDESeq2, a Python implementation of the DESeq2 statistical framework.^34^ Raw counts were normalized using the median-of-ratios method to account for differences in sequencing depth and RNA composition. Differential expression was modeled using negative binomial generalized linear models with gene-specific dispersion estimation.

Preservation condition was included as the explanatory variable, with pooled clinical controls serving as the reference group. Differential expression was evaluated using Wald tests of the null hypothesis that the log₂ fold change between preservation conditions was zero. Cook’s distance-based outlier handling and model refitting were enabled where supported by the available biological replicates. Resulting P values were adjusted for multiple comparisons using the Benjamini–Hochberg procedure to control the false discovery rate (FDR).

Genes with FDR <0.05 were considered statistically significant, with an additional absolute log₂ fold-change threshold >1 applied to identify differentially expressed genes exhibiting changes greater than twofold. Positive log₂ fold changes indicated increased expression in HOPE20h relative to clinical controls. Preranked gene set enrichment analysis (GSEA) was performed using GSEApy to identify coordinated transcriptional differences in predefined biological pathways.^35^ Genes were ranked according to their signed Wald test statistics obtained from PyDESeq2, thereby incorporating both the direction and statistical strength of differential expression. All eligible genes with finite ranking statistics were included, irrespective of individual gene-level statistical significance or fold-change magnitude. Enrichment analyses were performed using MSigDB Hallmark 2020 and Gene Ontology Biological Process 2023 gene set collections. GSEA was performed using 1,000 permutations, a minimum gene set size of 10, a maximum gene set size of 500, and a fixed random seed of 42. Enrichment scores were calculated according to the distribution of pathway-associated genes within the ranked dataset and normalized to generate normalized enrichment scores (NES). Statistical significance was assessed using permutation-derived nominal P values and FDR-adjusted q-values, with FDR <0.05 defining significant pathway enrichment. Positive NES values indicated relative enrichment in HOPE20h, whereas negative NES values indicated relative enrichment in pooled clinical preservation controls. Pathway enrichment was interpreted as evidence of coordinated transcriptional differences rather than direct functional activation or inhibition of the corresponding biological processes.

### Cell-Type-Associated Transcriptional Signature Analysis

To investigate preservation-associated alterations in cardiac cell-type transcriptional signatures, bulk RNA sequencing data from porcine myocardial samples subjected to static cold storage for 4 hours (SCS4h; n=2), hypothermic oxygenated perfusion for 4 hours (HOPE4h; n=2), or extended hypothermic oxygenated perfusion for 20 hours (HOPE20h; n=4) were analyzed using a custom Python-based pipeline. Raw gene-level counts were normalized using the median-of-ratios method, with size factors calculated from genes exhibiting nonzero counts across all samples. Normalized expression values were subsequently log₂-transformed following the addition of a pseudocount of one. To characterize cell-type-associated transcriptional activity, curated marker gene panels were assembled for cardiomyocytes, fibroblasts, endothelial cells, macrophages/monocytes, T lymphocytes, B lymphocytes, pericytes, and vascular smooth muscle cells (Supplemental Figure 10A).^36–39^ Candidate marker genes were mapped to porcine Ensemble gene identifiers using MyGene.info (Sus scrofa, NCBI taxonomy ID: 9823). Only exact gene-symbol matches with unambiguous gene-level assignments were retained. Marker coverage was assessed for each cell population, and signatures containing fewer than three successfully mapped, variable genes were excluded. To evaluate relative cell-type-associated transcriptional activity, log₂-transformed expression values were standardized independently for each gene across all eight biological samples using z-score normalization. For each sample, cell-type-associated transcriptional signature scores were calculated as the arithmetic mean of the standardized expression values of all successfully mapped marker genes within the corresponding cell-type panel. Importantly, differences in signature scores cannot distinguish altered cellular abundance from transcriptional regulation within existing cell populations. The limited number of biological replicates constrained the resolution and statistical power of exact permutation testing, particularly in comparisons involving two control samples.

### GSEA Leading-Edge Analysis and Pathway Redundancy Mapping

Leading-edge analysis was performed using a custom Python pipeline to identify genes contributing to pathway enrichment and characterize redundancy among enriched biological pathways. Leading-edge gene annotations, normalized enrichment scores (NES), and false discovery rate (FDR) q-values were obtained from the preceding gene set enrichment analysis (GSEA). Leading-edge genes were extracted from the GSEA results as the subset of pathway-associated genes contributing to the enrichment score at its maximum deviation from zero. These genes represent the principal contributors to the observed enrichment signal within each pathway. To assess redundancy among enriched biological pathways, pairwise overlap of leading-edge gene sets was quantified using the Jaccard similarity coefficient defined as the number of shared leading-edge genes divided by the total number of unique genes represented across both pathways. Gene–pathway relationships were additionally represented using binary membership matrices, indicating whether individual genes belonged to the leading-edge subset of each pathway. Membership heatmaps included up to 30 genes ranked by their frequency of occurrence across the top 25 significantly enriched pathways by minimum FDR. These matrices represented pathway membership rather than gene-expression magnitude. FDR in this analysis is pathway-level GSEA FDR, not gene-level DESeq2 FDR. A gene appearing in many leading edges is a recurring contributor to pathway enrichment, not automatically a significant DEG or a causal regulator. The Jaccard threshold (≥ 0.25) and gene-prioritization metrics were considered exploratory analytical criteria rather than independently validated statistical significance thresholds.

### Proteomics

Tissue biopsied from the IVS weighting approximately 200 mg was placed in cryotubes, snap-frozen in liquid nitrogen, and stored at −80°C until use.

#### Tissue Lysis and Protein Extraction

Samples were pulverized and suspended in lysis buffer containing 1% SDC, 2% SDS, and 1× protease inhibitor in 100 mM Tris-HCl (pH 8.0), followed by homogenization using PreOmics’ BeatBox for 10 min. The tubes were then placed on a magnetic rack, and the solubilized lysate was transferred to 1.5 mL Eppendorf tubes. The lysates were centrifuged at 14,000 rpm for 5 min, and the supernatants were transferred to fresh 1.5 mL Eppendorf tubes. The samples were incubated at 60 °C for 20 min with constant agitation at 1,400 rpm, followed by incubation at room temperature for 5 min and water-bath sonication for 2 min. Protein concentrations were determined using the Pierce BCA Protein Assay Kit.

A total of 10 µg of protein from each lysate was used for subsequent processing. Proteins were reduced and alkylated using 10 mM tris(2-carboxyethyl)phosphine (TCEP) and 40 mM 2-chloroacetamide at 45 °C for 10 min with constant agitation at 1,400 rpm. Following reduction and alkylation, the samples were incubated at room temperature for 5 min and sonicated in a water bath for 2 min. Lysate cleanup was performed using SP3 magnetic beads, followed by overnight digestion at 37 °C with Lys-C/Trypsin (Promega) at an enzyme-to-protein ratio of 1:100.^37^ Digestion was terminated by adding TFA to a final concentration of 1% (v/v). The resulting peptide mixtures were clarified by centrifugation at 14,000g for 10 min and desalted using a C18 plate.^41^ The eluted peptides were dried using a SpeedVac centrifuge and resuspended in 3% acetonitrile / 0.1% formic acid. An aliquot corresponding to 200 ng of peptides from each sample was subjected to LC-MS/MS analysis.

#### Liquid Chromatography - Tandem Mass Spectrometry (LC–MS/MS)

Peptides were separated over a 40-min gradient at a flow rate of 400 nl/min on a reverse-phase C18 column with an integrated CaptiveSpray emitter (25 cm × 75 µm, 1.6 µm; IonOpticks). Mobile phase A consisted of 0.1% formic acid in water, and mobile phase B consisted of 0.1% formic acid in acetonitrile. The gradient was programmed as follows: 2% to 25% B over 30 min, 25% to 40% B over 5 min, then increased to 95% B before column re-equilibration.

The timsTOF Pro2 was operated in diaPASEF model.^38^ Data were acquired using 60 variable isolation windows covering m/z 350-1,250. Collision energy was linearly ramped with ion mobility, ranging from 59 eV at 1/K₀ = 1.45 Vs cm⁻² to 20 eV at 1/K₀ = 0.6 Vs cm⁻².

#### Data Analysis

Raw diaPASEF files were searched against the UniProt Pig proteome database (UP000008227) using DIA-NN (v2.6.0) with default settings for library-free analysis ^39^. Search was performed with trypsin specificity, allowing up to one missed cleavage. Carbamidomethylation of cysteine (+57.021 Da) was set as a fixed modification, while protein N-terminal acetylation (+42.010 Da), methionine oxidation (+15.995 Da), and deamidation of N/Q (+0.984 Da) were included as variable modifications. The false discovery rate (FDR) was controlled at 1% at both precursor and protein levels. DIA-NN output files were further analyzed for differential abundance using DIA-Analyst platform (v0.11.1), without additional normalization. Missing values were imputed using the Perseus-type imputation method, and FDR was corrected with t-statistics based method. Proteins with adjusted p-value < 0.05 and absolute log2 fold change >1 were considered differentially abundant between groups. Gene set enrichment analysis (GSEA) was performed on quantitative proteomic data to identify biological pathways associated with extended hypothermic oxygenated heart preservation (HOPE20h) relative to pooled clinical preservation controls (SCS4h and HOPE4h). Preranked GSEA was conducted using GSEApy, with proteins ranked according to a signed statistical significance metric incorporating both the magnitude and direction of differential protein abundance.^44^ For each protein, the ranking statistic was calculated as: Ranking statistic = sign(log₂FC) × [−log₁₀(P value)], where log₂FC represented the relative protein abundance in HOPE20h compared with pooled clinical controls, and P represented the nominal P value obtained from differential protein abundance analysis. When nominal P values were unavailable, log₂ fold changes were used as the alternative ranking metric. Proteins with nonfinite ranking statistics were excluded, and duplicate gene symbols were removed to retain a single entry per gene. Preranked enrichment analysis was performed using MSigDB Hallmark 2020, Reactome 2022, and Gene Ontology Biological Process 2023 gene set collections. Gene sets were evaluated using 1,000 permutations, a minimum gene set size of 5, a maximum gene set size of 500, and a fixed random seed of 42. All eligible quantified proteins were included in the ranked dataset irrespective of their individual statistical significance.

Enrichment scores were calculated based on the distribution of pathway-associated proteins within the ranked dataset and normalized to generate normalized enrichment scores (NES). Statistical significance was assessed using permutation-derived nominal P values and false discovery rate (FDR)-adjusted q-values.

Pathways with FDR <0.05 were considered significantly enriched. Positive NES values indicated relative enrichment of pathway-associated proteins in HOPE20h, whereas negative NES values indicated relative enrichment in pooled clinical controls.

### Integrated Transcriptomic-Proteomics

RNA–protein directional concordance was evaluated using matched gene–protein pairs. A descriptive absolute log₂ fold-change threshold of 0.25 was applied independently to RNA and protein measurements.^45^ Pairs exceeding this threshold in both omics layers were classified as concordantly increased, concordantly decreased, RNA-increased/protein-decreased, or RNA-decreased/protein-increased according to their respective fold-change directions. Pairs failing to exceed the threshold in either layer were classified as below the directional threshold. Directional classifications were descriptive and independent of statistical significance, which was assessed separately using an FDR threshold of <0.05.

## List of Abbreviations

DBD: Donation after Brain Death
DCD: Donation after Circulatory Death
HOPE: Hypothermic Oxygenated Perfusion
SCS: Static Cold Storage
PGD: Primary Graft Dysfunction
CPB: Cardiopulmonary Bypass
HT: Heart Transplant
MCS: Mechanical Circulatory Support
NMP: Normothermic Machine Perfusion
HTK: Histidine-tryptophan-ketoglutarate

## References

1. Katz J, Waters S, Hollis I, Chang P. Advanced Therapies For End-Stage Heart Failure. Curr Cardiol Rev. 2014;11(1):63–72. doi:10.2174/1573403X09666131117163825

2. Bounader K, Flécher E. End-stage heart failure: The future of heart transplant and artificial heart. Presse Médicale. 2024;53(1):104191. doi:10.1016/j.lpm.2023.104191

3. Sundararaju U, Rachoori S, Mohammad A, Rajakumar HK. Cardiac transplantation: A review of current status and emerging innovations. World J Transplant. 2025;15(2). doi:10.5500/wjt.v15.i2.100460

4. Olsen BA, Miyamoto SD. Pediatric heart failure: Current approach and treatment. JHLT Open. 2025;9:100331. doi:10.1016/j.jhlto.2025.100331

5. Yazji JH, Garg P, Wadiwala I, et al. Expanding Selection Criteria to Repairable Diseased Hearts to Meet the Demand of Shortage of Donors in Heart Transplantation. Cureus. Published online May 30, 2022. doi:10.7759/cureus.25485

6. Toro S, Schlendorf KH. Expanding the Donor Pool. Methodist DeBakey Cardiovasc J. 2025;21(3):33–39. doi:10.14797/mdcvj.1577

7. Sponga S, Vendramin I, Salman J, et al. Heart Transplantation in High-Risk Recipients Employing Donor Marginal Grafts Preserved With Ex-Vivo Perfusion. Transpl Int. 2023;36:11089. doi:10.3389/ti.2023.11089

8. Song C, Iyengar A, Rekhtman D, et al. Use of extended criteria donor cardiac allografts after multiple offers is associated with inferior post-transplant outcomes. JHLT Open. 2026;13:100566. doi:10.1016/j.jhlto.2026.100566

9. Schroder JN, Patel CB, DeVore AD, et al. Transplantation Outcomes with Donor Hearts after Circulatory Death. N Engl J Med. 2023;388(23):2121–2131. doi:10.1056/NEJMoa2212438

10. Gaffey AC, Doll SL, Thomasson AM, et al. Transplantation of “high-risk” donor hearts: Implications for infection. J Thorac Cardiovasc Surg. 2016;152(1):213–220. doi:10.1016/j.jtcvs.2015.12.062

11. McGiffin DC, Kure CE, Macdonald PS, et al. Hypothermic oxygenated perfusion (HOPE) safely and effectively extends acceptable donor heart preservation times: Results of the Australian and New Zealand trial. J Heart Lung Transplant. 2024;43(3):485–495. doi:10.1016/j.healun.2023.10.020

12. Kosuri Y, Moroi MK, Bauer SJ, et al. Understanding donor heart preservation: Cellular mechanisms and clinical outcomes. Perfusion. 2026;41(5):547–558. doi:10.1177/02676591251377571

13. Critchley WR, Stone JP, Liao Q, et al. Non-ischemic Heart Preservation via Hypothermic Cardioplegic Perfusion Induces Immunodepletion of Donor Hearts Resulting in Diminished Graft Infiltration Following Transplantation. Front Immunol. 2020;11:1621. doi:10.3389/fimmu.2020.01621

14. Lu S, Karunasumetta C, Gao W, et al. Hypothermic perfusion protocol development for donor heart preservation. JTCVS Open. 2026;32:101845. doi:10.1016/j.xjon.2026.101845

15. Joshi Y, MacLean C, Emmanuel S, et al. Australian outcomes from heart transplantation in the machine perfusion era. Ann Cardiothorac Surg. 2024;13(6):502–512. doi:10.21037/acs-2024-dcd-0074

16. Hettlich VH, Aubin H, Scherner M, Ramadani B, Lichtenberg A, Boeken U. New Perfusion and Preservation Strategies for Donor Hearts. Eur Surg Res. 2026;67(1):99–106. doi:10.1159/000551946

17. Schlegel A, Muller X, Mueller M, et al. Hypothermic oxygenated perfusion protects from mitochondrial injury before liver transplantation. EBioMedicine. 2020;60:103014. doi:10.1016/j.ebiom.2020.103014

18. Rega F, Lebreton G, Para M, et al. Hypothermic oxygenated perfusion of the donor heart in heart transplantation: the short-term outcome from a randomised, controlled, open-label, multicentre clinical trial. The Lancet. 2024;404(10453):670–682. doi:10.1016/S0140-6736(24)01078-X

19. Camillo C, Moroi MK, Kosuri Y, et al. Ex Vivo Hypothermic Perfusion Enables 48-Hour Heart Preservation and Bench-Top Functional Recovery via Normothermic Reperfusion in a Porcine Model. J Am Heart Assoc. 2026;15(7):e045800. doi:10.1161/JAHA.125.045800

20. Moroi MK, Kosuri Y, Karcher C, et al. Molecular and cellular adaptations to extended hypothermic oxygenated perfusion in donation-after-circulatory-death hearts in a porcine model. Front Cardiovasc Med. 2026;13:1800470. doi:10.3389/fcvm.2026.1800470

21. Thomassen MB, Ballan EM, Moeslund N, et al. Twenty-four–hour hypothermic oxygenated perfusion preserves graft viability in a porcine donation after circulatory death and donation after brain death heart transplant model. J Heart Lung Transplant. 2026;45(7):1135–1146. doi:10.1016/j.healun.2026.03.021

22. Lionetti V, Romano SL, Bianchi G, et al. Impact of Acute Changes of Left Ventricular Contractility on the Transvalvular Impedance: Validation Study by Pressure-Volume Loop Analysis in Healthy Pigs. Moretti C, ed. PLoS ONE. 2013;8(11):e80591. doi:10.1371/journal.pone.0080591

23. Schmidt CA, Fisher-Wellman KH, Neufer PD. From OCR and ECAR to energy: Perspectives on the design and interpretation of bioenergetics studies. J Biol Chem. 2021;297(4):101140. doi:10.1016/j.jbc.2021.101140

24. Bottermann K, Spychala A, Eliacik A, et al. Extracellular flux analysis in intact cardiac tissue slices—A novel tool to investigate cardiac substrate metabolism in mouse myocardium. Acta Physiol. 2023;239(2):e14004. doi:10.1111/apha.14004

25. Sciaccaluga C, Fusi C, Barilli M, et al. Diastolic function in heart transplant: From physiology to echocardiographic assessment and prognosis. Front Cardiovasc Med. 2022;9:969270. doi:10.3389/fcvm.2022.969270

26. Ettrup KS, Glud AN, Orlowski D, et al. Basic Surgical Techniques in the Göttingen Minipig: Intubation, Bladder Catheterization, Femoral Vessel Catheterization, and Transcardial Perfusion. J Vis Exp. 2011;(52):2652. doi:10.3791/2652

27. Hadaya J, Buckley U, Gurel NZ, et al. Scalable and reversible axonal neuromodulation of the sympathetic chain for cardiac control. Am J Physiol-Heart Circ Physiol. 2022;322(1):H105–H115. doi:10.1152/ajpheart.00568.2021

28. Vrabec T, Bender S, Chan S, et al. Bioelectronic block of stellate ganglia mitigates pacing-induced heterogeneous release of catecholamine and neuropeptide Y in the infarcted pig heart. J Physiol. 2025;603(7):2071–2088. doi:10.1113/JP286924

29. Paccalet A, Tessier N, Paillard M, et al. An innovative sequence of hypoxia-reoxygenation on adult mouse cardiomyocytes in suspension to perform multilabeling analysis by flow cytometry. Am J Physiol-Cell Physiol. 2020;318(2):C439–C447. doi:10.1152/ajpcell.00393.2019

30. Li Y, Li Y, Li Y, et al. MTX2 facilitates PKM2 tetramerization to promote cardiac glucose metabolism and protects the heart against ischemia/reperfusion injury. Theranostics. 2025;15(14):6737–6752. doi:10.7150/thno.110162

31. Hu T, Wu Q, Yao Q, et al. PRDM16 exerts critical role in myocardial metabolism and energetics in type 2 diabetes induced cardiomyopathy. Metabolism. 2023;146:155658. doi:10.1016/j.metabol.2023.155658

32. Bottermann K, Spychala A, Eliacik A, et al. Extracellular flux analysis in intact cardiac tissue slices—A novel tool to investigate cardiac substrate metabolism in mouse myocardium. Acta Physiol. 2023;239(2):e14004. doi:10.1111/apha.14004

33. Mdaki KS, Larsen TD, Weaver LJ, Michelle L. Baack. Age Related Bioenergetics Profiles in Isolated Rat Cardiomyocytes Using Extracellular Flux Analyses. Akar FG, ed. PLOS ONE. 2016;11(2):e0149002. doi:10.1371/journal.pone.0149002

34. Muzellec B, Teleńczuk M, Cabeli V, Andreux M. PyDESeq2: a python package for bulk RNA-seq differential expression analysis. Ponty Y, ed. Bioinformatics. 2023;39(9):btad547. doi:10.1093/bioinformatics/btad547

35. Fang Z, Liu X, Peltz G. GSEApy: a comprehensive package for performing gene set enrichment analysis in Python. Lu Z, ed. Bioinformatics. 2023;39(1):btac757. doi:10.1093/bioinformatics/btac757

36. Litviňuková M, Talavera-López C, Maatz H, et al. Cells of the adult human heart. Nature. 2020;588(7838):466–472. doi:10.1038/s41586-020-2797-4

37. Tucker NR, Chaffin M, Fleming SJ, et al. Transcriptional and Cellular Diversity of the Human Heart. Circulation. 2020;142(5):466–482. doi:10.1161/CIRCULATIONAHA.119.045401

38. Chaffin M, Papangeli I, Simonson B, et al. Single-nucleus profiling of human dilated and hypertrophic cardiomyopathy. Nature. 2022;608(7921):174–180. doi:10.1038/s41586-022-04817-8

39. The Tabula Sapiens Consortium*, Jones RC, Karkanias J, et al. The Tabula Sapiens: A multiple-organ, single-cell transcriptomic atlas of humans. Science. 2022;376(6594):eabl4896. doi:10.1126/science.abl4896

40. Soni RK. Protocol for deep proteomic profiling of formalin-fixed paraffin-embedded specimens using a spectral library-free approach. STAR Protoc. 2023;4(3):102381. doi:10.1016/j.xpro.2023.102381

41. Soni RK. High-Throughput Plasma Proteomic Profiling. In: Garg U, ed. Clinical Applications of Mass Spectrometry in Biomolecular Analysis. Vol 2546. Methods in Molecular Biology. Springer US; 2022:411–420. doi:10.1007/978-1-0716-2565-1_36

42. Meier F, Brunner AD, Frank M, et al. diaPASEF: parallel accumulation–serial fragmentation combined with data-independent acquisition. Nat Methods. 2020;17(12):1229–1236. doi:10.1038/s41592-020-00998-0

43. Demichev V, Messner CB, Vernardis SI, Lilley KS, Ralser M. DIA-NN: neural networks and interference correction enable deep proteome coverage in high throughput. Nat Methods. 2020;17(1):41–44. doi:10.1038/s41592-019-0638-x

44. Subramanian A, Tamayo P, Mootha VK, et al. Gene set enrichment analysis: A knowledge-based approach for interpreting genome-wide expression profiles. Proc Natl Acad Sci. 2005;102(43):15545–15550. doi:10.1073/pnas.0506580102

45. Shen X, Hu Q, Li J, Wang J, Qu J. Experimental Null Method to Guide the Development of Technical Procedures and to Control False-Positive Discovery in Quantitative Proteomics. J Proteome Res. 2015;14(10):4147–4157. doi:10.1021/acs.jproteome.5b00200

